# Venetoclax-based Therapy Improves Outcomes across the Evolving Biology of t(11;14) Multiple Myeloma

**DOI:** 10.64898/2026.09.02.26361956

**Authors:** Praneeth Reddy Sudalagunta, Filip Ionescu, Rafael Renatino Canevarolo, Maria Coelho Siqueira Silva, Daniel DeAvila, Mark B. Meads, Xiaohong Zhao, Angel Perez, Dimitrios Drekolias, Ruxandra Irimia, Shrinjaya Thapa, Parth Patel, Rachid Baz, Kenneth H. Shain, Ariosto Siqueira Silva, Ariel Grajales-Cruz

## Abstract

**Background:** Translocation t(11;14) defines a biologically distinct subset of multiple myeloma (MM) enriched for BCL2 dependency. Treatment with venetoclax, selective BCL2 inhibitor, has shown varied responses in clinical trials and retrospective cohorts. Efficacy of venetoclax-based combination therapies and optimal timing of treatment in t(11;14) MM patients remain incompletely characterized.

**Methods:** We have compared the overall survival of t(11;14) MM patients who received Venetoclax (N = 97) at any moment in time, to those who never did (N =284), in the largest retrospective cohort (N = 381) reported to date. We used longitudinal fluorescence in situ hybridization (FISH) to assess cytogenetic evolution and genomic complexity. We performed transcriptomic profiling of CD138-enriched tumors using RNA sequencing in a subset of samples and gene-expression signatures (UAMS/HALLMARKS) were used to define molecular subtypes. Ex vivo drug sensitivity assays integrated with paired RNA sequencing were used to identify subtype-specific therapeutic vulnerabilities and rational venetoclax-based combination strategies.

**Results:** Venetoclax exposure was associated with an improvement of median overall survival by nearly four years compared to non–VEN-exposed patients (p = 0.0003). Longitudinal cytogenetic analysis demonstrated stability of the primary t(11;14) translocation over time, while secondary abnormalities tend to accumulate, including those harboring high-risk secondary cytogenetic abnormalities such as del13q, amp/gain1q21, del17p, and del1p, resulting in increasing genomic complexity with disease progression. Transcriptomic analyses identified selective enrichment of CD1/CD2 signature (associated with t(11;14) NDMM) by single-sample gene set enrichment analysis as a marker of prolonged progression-free survival. However, patients with more than five prior lines of therapy were enriched for transcriptionally complex biology characterized by CD1/CD2 with either proliferative/hypermetabolic or inflammatory transcriptional programming which were associated with inferior outcomes with VEN-based treatment. Ex vivo drug sensitivity profiling revealed subtype-specific vulnerabilities, identifying daratumumab, lenalidomide, ixazomib, and panobinostat as rational partners for venetoclax depending on transcriptional context.

**Conclusion:** Venetoclax-based therapy significantly improved overall survival in t(11;14) MM. Clinical benefit, defined by improved progression free survival, was greatest when venetoclax was administered earlier, preceding the emergence of transcriptomic reprogramming that reduces BCL2 dependency. These findings support transcriptomic biomarker-guided, subtype-specific venetoclax-based treatment strategies to optimize outcomes in patients with t(11;14) MM.

## Background

The treatment of multiple myeloma (MM), the second most prevalent hematologic malignancy, has seen remarkable clinical progress with the introduction of therapies directed against diverse aspects of plasma cell biology^1^. Despite these advances, the development and use of truly targeted therapies have lagged behind^2,3^. Broadly, MM is characterized by primary cytogenetic abnormalities which consist of hyperdiploidy involving trisomy of odd-numbered chromosomes or translocations of the immunoglobulin heavy chain (*IGH*) locus at 14q32 with various oncogenes^4^. Among the latter, the t(11;14)(q13;q32) translocation, which places the *IGH* enhancer in proximity to *CCND1*, represents a distinct biological subgroup with unique clinical and therapeutic implications. The t(11;14) translocation, originally prognostically neutral, has now been associated with inferior outcomes in the era of novel therapies. Immunomodulatory drugs (IMiDs), proteasome inhibitors (PIs), and monoclonal antibodies (mAbs) preferentially benefit non-t(11;14) patients and place those with the translocation in an intermediate-risk group between standard- and high-risk disease^5,6,7^. In the recent MIDAS trial (NCT04934475) involving transplant-eligible newly diagnosed MM (NDMM) patients treated with a novel four-drug combination (isatuximab, carfilzomib, lenalidomide, and dexamethasone) for six cycles as induction therapy, only 24% of t(11;14) MM patients achieved minimal residual disease (MRD) negativity compared to 59% of non-t(11;14) MM patients^8^. These findings highlight the need for new therapeutic strategies specifically designed to target the unique biology of t(11;14) MM in the clinic.

t(11;14) MM is characterized by overexpression of *CCND1* and a lymphoid/B cell-like transcriptional program, which is associated with overexpression of *BCL2*^9^. This increase in *BCL2* expression corresponds with upregulation of *PMAIP1* (*NOXA*) and *BBC3* (*PUMA*), and higher ratios of *BCL2*/*MCL1* and *BCL2*/*BCL2L1* (*BCL-xL*) expression^10^. NOXA and PUMA bind with high affinity to MCL1 and BCL-xL, which leaves BCL2 to sequester BIM to maintain cell survival^11^. This BCL2-dependence can be therapeutically exploited via venetoclax (VEN), a BH3-mimetic BCL2 inhibitor, driving apoptosis^12,13^. The efficacy and tolerability of VEN have been evaluated in multiple clinical trials and retrospective patient cohorts. In the phase I/II trial^14^ involving VEN monotherapy, t(11;14) MM patients showed deeper responses and prolonged progression-free survival (PFS) with acceptable tolerability. Moreover, phase III CANOVA trial (NCT03539744)^15,16^, phase I/II trial (NCT03314181)^17,18^, phase II trial (NCT02899052)^19^, and phase III BELLINI trial (NCT02755597)^20,21^ showed encouraging results in t(11;14) MM for VEN treatment when combined with IMiDs, anti-CD38 mAbs, and proteasome inhibitors, respectively. In these clinical trials, VEN-based regimens (combined with standard-of-care) consistently yielded deeper responses in t(11;14) MM patients improving ORR compared to t(11;14) MM patients treated with standard-of-care regimens, however, PFS benefit varied considerably. Subsequent real-world, off-label use of VEN in more heavily pretreated patients with t(11;14) MM has shown a general trend toward decreasing progression-free survival (PFS) with increasing prior lines of therapy (LOT), with median PFS reported as 12 months after a median of 3 prior LOT^22,23^, 10 months (4 median prior LOT)^24^, and 7.8 months (5 median prior LOT)^25^. An outlier study reported a median PFS of 11.2 months after 7 prior LOT^26^.

Although patients with t(11;14) MM harbor a clear biological dependency on BCL2 that can be therapeutically targeted with VEN, existing studies have not yet translated this vulnerability into consistently durable clinical benefit. Key questions remain about the integration of VEN in the management of this subgroup, such as improved patient selection, optimal sequencing and preferred partner agents that most effectively enhance VEN efficacy in t(11;14) MM patients. To address these issues, we evaluated VEN efficacy in relation to concurrent cytogenetic abnormalities, partner drugs, gene expression profiles, and ex vivo drug sensitivity in the largest real-world cohort of t(11;14) MM reported to date.

## Methods

### Study design and cohort

We retrospectively identified all patients with a diagnosis of MM, who received treatment at Moffitt Cancer Center (Tampa, FL) between September 2000 and June 2023, and a positive indication for t(11;14) by fluorescence in situ hybridization (FISH) assay any time after diagnosis. Patients with primary plasma cell leukemia, concurrent systemic amyloidosis, active solid malignancies, or concurrent hematologic malignancies were excluded. These inclusion/exclusion criteria resulted in 381 patients. Of these, 97 received VEN any time after MM diagnosis (VEN-exposed) and 284 did not (Not VEN-exposed). No significant differences in demographic characteristics (age, biological sex, and race) were noted between the two groups. The following variables were populated from clinical notes: ISS staging, extramedullary disease status at MM diagnosis, type of myeloma (intact immunoglobulin, light chain, and oligo or non-secretory), cytogenetic abnormalities by FISH – amp/gain1q21, del1p, del13q, del17p, t(4;14), t(11;14), and t(14;16), lines of treatment (LOT) at follow-up and prior LOT for VEN-exposed only, and exposure to standard-of-care therapeutic interventions (lenalidomide, pomalidomide, bortezomib, carfilzomib, anti-CD38 antibody, penta-exposed, autologous stem cell transplant – ASCT, CAR-T cell therapy, and bispecific antibodies).

The medical records were de-identified and only the following clinically relevant information was reviewed: (A) the treatment administered (therapeutic agents, doses, and schedule) following the biopsy, (B) cytogenetics, (C) disease statuses, (D) demographics, and (E) treatment outcomes. Patient samples were used in accordance with the Declaration of Helsinki, International Ethical Guidelines for Biomedical Research Involving Human Subjects (CIOMS), Belmont Report, and U.S. Common Rule.

### FISH-based genonic conplexity

The cytogenetic status was evaluated using CLIA-certified FISH assays profiling CD138-enriched tumor samples for the presence of (or lack thereof) t(11;14) (>0.5% is positive), t(4;14) (>0.5% is positive), t(14;16) (>0.5% is positive), del1p (>3% is positive), amp/dup1q21 (>3% is positive), del13q (>4% is positive), and del17p (>4.5% is positive). The 381-patient cohort for our study was informed by the t(11;14)-positive status, where secondary cytogenetic abnormalities amp/dup1q21, del13q, and del17p were considered to inform genomic complexity of the sample/patient. At(11;14)-simple status was defined by t(11;14)-positivity and negative status for all secondary cytogenetic abnormalities. First-order, second-order, and third-order complexity were defined by the presence of one, two, or three secondary cytogenetic abnormalities, respectively, in addition to t(11;14).

### RNA sequencing

Fresh BM aspirate cells were enriched for CD138 expression using Miltenyi (Bergisch Gladbach, Germany) 130-051-301 antibody-conjugated magnetic beads. 1.0 x 10^6^ viably frozen CD138+ cells were shipped for molecular analysis in the context of the ORIEN AVATAR program. For frozen tissue RNA extraction, Qiagen RNAeasy plus mini kit was performed, generating 216 bp average insert size. RNA Sequencing was performed using the Illumina TruSeq RNA Exome with single library hybridization, cDNA synthesis, library preparation, sequencing (at either 100 or 150 bp paired reads) to a coverage of 100M total reads / 50M paired reads. RNA-seq tumor pipeline analysis was processed according to the workflow outlined below using GRCh38/hg38 human genome reference sequencing and GenCode build version 32.

#### Adapter trimming

Adapter sequences were trimmed from the raw tumor sequencing FASTQ file. Adapter-trimming via k-mer matching was performed along with quality-trimming and filtering, contaminant-filtering, sequence masking, GC-filtering, length filtering and entropy-filtering. The trimmed FASTQ file was used as input to the read alignment process.

#### Read Alignment

The tumor adapter-trimmed FASTQ file was aligned to the human genome reference (GRCh38/hg38) and the Gencode genome annotation v32 using the STAR aligner. The STAR aligner generates multiple output files used for Gene Fusion Prediction and Gene Expression Analysis.

#### RNA expression

RNA expression values were calculated and reported using estimated mapped reads, Fragments Per Kilobase of transcript per Million mapped reads (FPKM), and Transcripts Per Million mapped reads (TPM) at both transcript level and gene level based on transcriptome alignment generated by STAR. Gene expression data was obtained from DNAnexus files containing FPKM and TPM values for 59,368 records. Among these, 19,933 were protein coding genes, which were further analyzed; the remainder were discarded. For each gene/sample, we calculated log2(FPKM+10^-3^) and removed any genes whose values for quartile 1 and quartile 3 were the same (i.e., any gene must be expressed in at least 25% of samples to be considered in this analysis). The remaining 16,738 genes were z-normalized across all samples using MATLAB’s function normalize.

### Transcriptonic subtyping

Single-sample gene set enrichment analysis (ssGSEA, RRID:SCR_026610) was used for transcriptomic subtyping of CD138-enriched tumor samples using their gene expression profiles from RNA sequencing. We relied on the gene set definitions available in msigdb (Molecular Signatures Database, RRID:SCR_016863) for each of the molecular subtypes identified in Zhan et al. (CD1_AND_CD2_UP, CD1_AND_CD2_DN, CD1_UP, CD1_DN, CD2_UP, CD2_DN, MS_UP, MS_DN, MF_UP, MF_DN, PR_UP, PR_DN, LB_UP, and LB_DN)^27^. Based on the msigdb gene set definitions, we computed ssGSEA scores and their corresponding statistical significance using false discovery rate (FDR) estimates for each t(11;14) MM RNAseq sample and every gene set. For every non-significant ssGSEA score (FDR > 0.05), we assign a zero-enrichment score (to not account for non-significant enrichment) and subtract the DN gene set score from the UP gene set score (to compute a unified enrichment score) for each of CD1 & CD2, CD1, CD2, MS, MF, PR, and LB gene sets. An ideal enrichment for a gene set would require a statistically significant positive enrichment score for the UP-gene set and a statistically significant negative enrichment score for the DN-gene set. These unified and statistically rigorous enrichment scores were used for transcriptomic subtyping of t(11;14) MM RNAseq samples, where all t(11;14) MM samples had positive scores for CD1 & CD2, while some were positive for CD1 and others were positive for CD2. In addition to a positive CD1 & CD2, a subset of t(11;14) MM samples also had a positive MF/PR score (and negative or zero for LB), while other samples had a positive LB score regardless of their MF/PR status.

### Ex vivo drug sensitivity assays

An ex vivo assay was used to quantify the chemosensitivity of primary MM cells. Fresh BM aspirate cells were enriched for CD138+ expression using Miltenyi (Bergisch Gladbach, Germany) 130-051-301 antibody-conjugated magnetic beads. MM cells (CD138+) were seeded in Corning (Corning, NY) CellBIND 384 well plates with collagen I and previously established human-derived stroma, containing approximately 4,000 MM cells and 1,000 stromal cells. Each well was filled with 80μL of RPMI-1640 media supplemented with fetal bovine serum (FBS, heat inactivated), penicillin/streptomycin, and patient-derived plasma (10%, freshly obtained from patient’s own aspirate, filtered) and left overnight for adhesion of stroma. The next day, drugs were added using a robotic plate handler so that every drug/combination was tested at 5 (fixed concentration ratio, for combinations) concentrations (1:3 serial dilution) in two replicates. Negative controls (supplemented growth media with and without the vehicle control dimethyl sulfoxide [DMSO]) were included, as well as positive controls for each drug (cell line MM1.S at highest drug concentration). Plates were placed in a motorized stage microscope (EVOS Auto FL, Life Technologies) equipped with an incubator and maintained at 5% CO2 and 37 °C. Each well was imaged every 30 min for a total duration of up to 6 days. A digital image analysis algorithm was implemented to determine changes in viability of each well longitudinally across 96h. This algorithm computes differences in sequential images and identifies live cells with continuous membrane deformations resulting from their interaction with the surrounding extracellular matrix. These interactions cease upon cell death. By applying this operation to all 288 images acquired for each well, we quantified non-destructively, and without the need to separate the stroma and myeloma, the effect of drugs as a function of concentration and exposure time. Digital image analysis computes percent viability of MM cells for each time point and experimental condition (drug and concentration). For each patient-drug, we have a dose-time-response surface, which is abstracted into AUC (area under the curve). The AUC is an area/integral measure of ex vivo response to therapy computed by taking an average of all ex vivo responses across all time (first 96h) and concentration. These patient-drug specific AUCs were used as representative measures of patient-specific response to each drug tested in ex vivo conditions.

### Predictive nachine learning nodels

#### Cytogenetic Data as Input

We developed a regression tree model to predict VEN ex vivo response using cytogenetic abnormalities detected by fluorescence in situ hybridization (FISH). The dataset included paired FISH and ex vivo AUC data from t(11;14) MM patients in the Moffitt cohort. Each FISH abnormality was encoded as a binary input variable representing the presence or absence of: t(11;14), t(4;14), t(14;16), del(1p), amp/dup(1q21), del(13q), and del(17p). The model was implemented in MATLAB R2022b using the *fitrtree* function from the *Statistics and Machine Learning Toolbox*. Inputs were patient-level binary vectors corresponding to these seven cytogenetic features, and the output was the continuous ex vivo VEN AUC. The trained regression tree generated a predicted ex vivo AUC for each patient using FISH data only. Each patient in the validation cohort subsequently received VEN as their immediate next line of therapy following the biopsy that produced the FISH results. To connect these model predictions with clinical outcome, we systematically evaluated predicted AUC thresholds (step size = 0.05) to stratify patients into predicted sensitive and predicted resistant groups. For each threshold, we performed Kaplan–Meier analysis of time-to-next-treatment (TTNT) on VEN and calculated log-rank p-values. The optimal predicted AUC cut-off was defined as the threshold yielding the lowest p-value, representing the point at which predicted ex vivo response most effectively discriminated TTNT between the two groups. This approach provided a quantitative FISH-based surrogate for VEN ex-vivo sensitivity.

#### RNAseq data as Input

We constructed a regression tree model to estimate VEN ex vivo AUC from transcriptomic features derived from RNA-seq data. Z-normalized expression profiles of 16,738 genes from 844 MM patients were analyzed, treating patients as variables defining a high-dimensional heterogeneity space and genes as observations. Dimensionality reduction using *t*-distributed Stochastic Neighbor Embedding (t-SNE) projected genes into a two-dimensional manifold preserving co-expression relationships. Genes in this embedded space were clustered using fuzzy C-means (FCM) to identify modules of co-expressing genes, yielding 500 clusters of varying sizes. Each cluster was tested for association with VEN response using Gene Set Enrichment Analysis (GSEA), ranking genes by their correlation with ex vivo AUC. Enrichment scores (ES) were computed using the running-sum statistic, and significance was determined via 1,000 phenotype permutations to generate null ES distributions. Clusters with FWER < 0.05 were retained as significantly enriched for sensitivity or resistance, defining the transcriptomic footprint of VEN in MM. For each statistically significant cluster, the median gene expression across its member genes was computed per patient, generating a matrix of median expression values representing module-level features. These values served as inputs to a regression tree model (*fitrtree*, MATLAB) and trained using ex vivo VEN AUC as the output variable. The resulting model predicted ex vivo AUC for each patient from RNA-seq data alone. Each patient then received VEN as their immediate next treatment following the RNA-seq biopsy. To identify the molecular threshold linked to clinical benefit, we scanned across predicted AUC values (step size = 0.05), stratified patients by predicted sensitivity, and performed Kaplan–Meier analysis of TTNT on VEN. The cut-off with the lowest log-rank p-value defined the predicted AUC that most effectively separated durable responders from early progressors, thereby establishing a transcriptomic surrogate of VEN sensitivity.

### Statistical analysis

#### Survival Analyses

Overall survival (OS) was defined from MM diagnosis to death or last follow-up. Time-to-next-treatment (TTNT) was defined from initiation of VEN to next therapy or death. Survival distributions were estimated using the Kaplan–Meier method, and group comparisons were made with the log-rank test. For cytogenetic evolution, the time from MM diagnosis to observation of cytogenetic complexity (gain of del13q, amp/gain1q21, del17p, or del1p) was analyzed using Kaplan–Meier curves and log-rank tests. Sequential FISH data were used to classify patients as showing *gain*, *loss*, or *no change* in complexity over time, and differences were compared similarly. Median survival and 95% confidence intervals (CIs) were estimated.

#### Forest Plots

Associations with OS and TTNT were assessed using reporting hazard ratios (HRs) with 95% CIs derived from univariable Cox proportional hazards models, where the presence of each variable was compared with the rest of the cohort. Variables included cytogenetic features (t(11;14) only, t(11;14)+del1p, t(11;14)+amp/gain1q21, t(11;14)+del13q, t(11;14)+del17p), molecular markers (e.g., BCL2-family markers and ratios), and partner drug category. Depth of response was modeled using binary logistic regression, where partial response or worse (≤PR was the dependent variable. Odds ratios (ORs) with 95% CIs were reported for cytogenetic abnormalities, partner drug type, and molecular markers ( e.g., BCL2-family markers and ratios). Forest plots were generated for OS, TTNT, and ≤PR analyses using HRs or ORs with corresponding CIs on a log₂ scale.

#### Group Comparisons

Baseline clinical and cytogenetic characteristics of VEN-exposed (n = 97) and Not VEN-exposed (n = 284) patients were compared. Continuous variables (e.g., age, number of prior lines) were tested using Wilcoxon rank-sum, and categorical variables (e.g., ISS stage, EMD, cytogenetic abnormalities, prior therapies) using Chi-square or Fisher’s exact test. Variables included age, sex, race, ISS stage, myeloma type, cytogenetics (t(11;14), t(4;14), t(14;16), del13q, amp/gain1q21, del17p, del1p), prior use of PIs, IMiDs, anti-CD38 antibodies, ASCT, CAR-T, and bispecific antibodies. Significant differences were highlighted in Table 1 and used for subgroup survival analyses in Supplementary Fig. 1.

**Table 1.** Demographic and disease-related features of t(11; 14)-positive mutiple myeloma patients.

| **Table 1. Demographic and disease-related features of t(11;14)-positive multiple myeloma patients** |  |  |  |  |
| --- | --- | --- | --- | --- |
|  | All patients<br>(N = 381) | Not VEN-exposed<br>(N = 284) | VEN-exposed<br>(N = 97) | P-value |
| **Age at diagnosis, years** |  |  |  |  |
| < 65 | 187 (49%) | 141 (50%) | 53 (54%) | .60 |
| ≥ 65 | 194 (51%) | 143 (50%) | 44 (45%) |  |
| Median (range) | 65 (33-86) | 65 (33-86) | 63 (34-81) | .22 |
| **Sex, male** | 236 (62%) | 175 (62%) | 61 (63%) | .90 |
| **Race** |  |  |  |  |
| White | 317 (83%) | 237 (83%) | 80 (82%) | .81 |
| Black | 40 (10%) | 28 (10%) | 12 (12%) |  |
| Other | 24 (7%) | 19 (10%) | 5 (6%) |  |
| **ISS disease stage** |  |  |  |  |
| I | 94 (25%) | 80 (28%) | 14 (14%) | - |
| II | 88 (23%) | 62 (22%) | 26 (27%) |  |
| III | 53 (14%) | 37 (13%) | 16 (16%) |  |
| Unknown | 146 (38%) | 105 (37%) | 41 (42%) |  |
| **Extramedullary disease at diagnosis** | 65 (17%) | 47 (17%) | 18 (19%) | .65 |
| **Myeloma type** |  |  |  |  |
| Intact immunoglobulin | 276 (72%) | 211 (74%) | 65 (67%) | .33 |
| Light chain only | 100 (26%) | 69 (24%) | 31 (32%) |  |
| Oligo-/nonsecretory | 5 (1%) | 4 (1%) | 1 (1%) |  |
| **Cytogenetic abnormalities at any time point** |  |  |  |  |
| High-risk abnormalities* | 58 (15%) | 44 (15%) | 21 (22%) | .35 |
| Del(17p) | 57 (15%) | 38 (13%) | 19 (20%) | .24 |
| t(4;14) | 2 (2%) | 0 (0%) | 2 (2%) | .06 |
| t(14;16) | 0 (0%) | 0 (0%) | 0 (0%) | .64 |
| gain(1q) or amp(1q) | 119 (31%) | 82 (29%) | 37 (38%) | .19 |
| Unknown | 3 (1%) | 2 (1%) | 1 (1%) | - |
| **Treatment exposure history** |  |  |  |  |
| Lines of treatment (median, range) | 3 (1-15) | 2 (1-13) | 5 (1-15) | <.01 |
| Lenalidomide | 143 (38%) | 95 (33%) | 48 (49%) | <.01 |
| Pomalidomide | 76 (20%) | 42 (15%) | 34 (35%) | <.01 |
| Bortezomib | 132 (35%) | 87 (31%) | 45 (46%) | <.01 |
| Carfilzomib | 71 (19%) | 39 (14%) | 32 (33%) | <.01 |
| Anti-CD38 antibody | 127 (33%) | 81 (29%) | 46 (47%) | <.01 |
| Penta-exposed | 50 (13%) | 25 (9%) | 25 (26%) | <.01 |
| ASCT | 196 (53%) | 134 (49%) | 62 (65%) | <.01 |
| CAR T-cell therapy | 48 (13%) | 10 (4%) | 38 (39%) | <.01 |
| Bispecific antibodies | 9 (2%) | 0 (0%) | 9 (9%) | <.01 |
| High-risk cytogenetics include del(17p), t(4;14), and t(14;16). Penta-exposed disease: exposed to lenalidomide, pomalidomide, bortezomib, carfilzomib, and daratumumab or isatuximab. |  |  |  |  |
| Abbreviations: ASCT, autologous stem cell transplant; ECOG PS, Eastern Cooperative Oncology Group performance status; ISS, Revised International Staging System; IMiD, immunomodulatory drugs; PI, proteasome inhibitor. |  |  |  |  |

## Results

### Exposure to Venetoclax Improves Overall Survival of t(11;14) Multiple Myeloma Patients

We have identified, between September 2000 and June 2023, 381 patients with t(11;14) MM treated at Moffitt Cancer Center; of these, 97 were treated with VEN at some point in time following MM diagnosis (Fig. 1A). No significant differences in association were observed between VEN-exposed and Not-VEN-exposed groups by their age, biological sex, race, International Staging System (ISS) status, Extramedullary Disease (EMD) status, type of MM (heavy chain/light chain/non-secretory), and incidence of cytogenetic abnormalities at any time since MM diagnosis (Table 1). Notably, the VEN-exposed group had a higher proportion of penta-exposed patients and a higher number of median LOT (5 vs 2 for the Not VEN-exposed group) at follow-up. Despite the diagnosis date falling within similar ranges (2003-2022 for VEN-exposed and 2000-2023 for Not-VEN-exposed), the VEN-exposed group had a higher proportion of patients treated with contemporary agents: BTZ (46% vs 31%), CFZ (33% vs 19%), POM (35% vs 15%), LEN (49% vs 33%), anti-CD38 mAb (47% vs 29%), as well as novel immune therapies such as chimeric antigen receptor T cell therapy (CAR-T, 39% vs 4%) and bispecific antibodies (9% vs 0%) (Table 1).

**Fig. 1.**
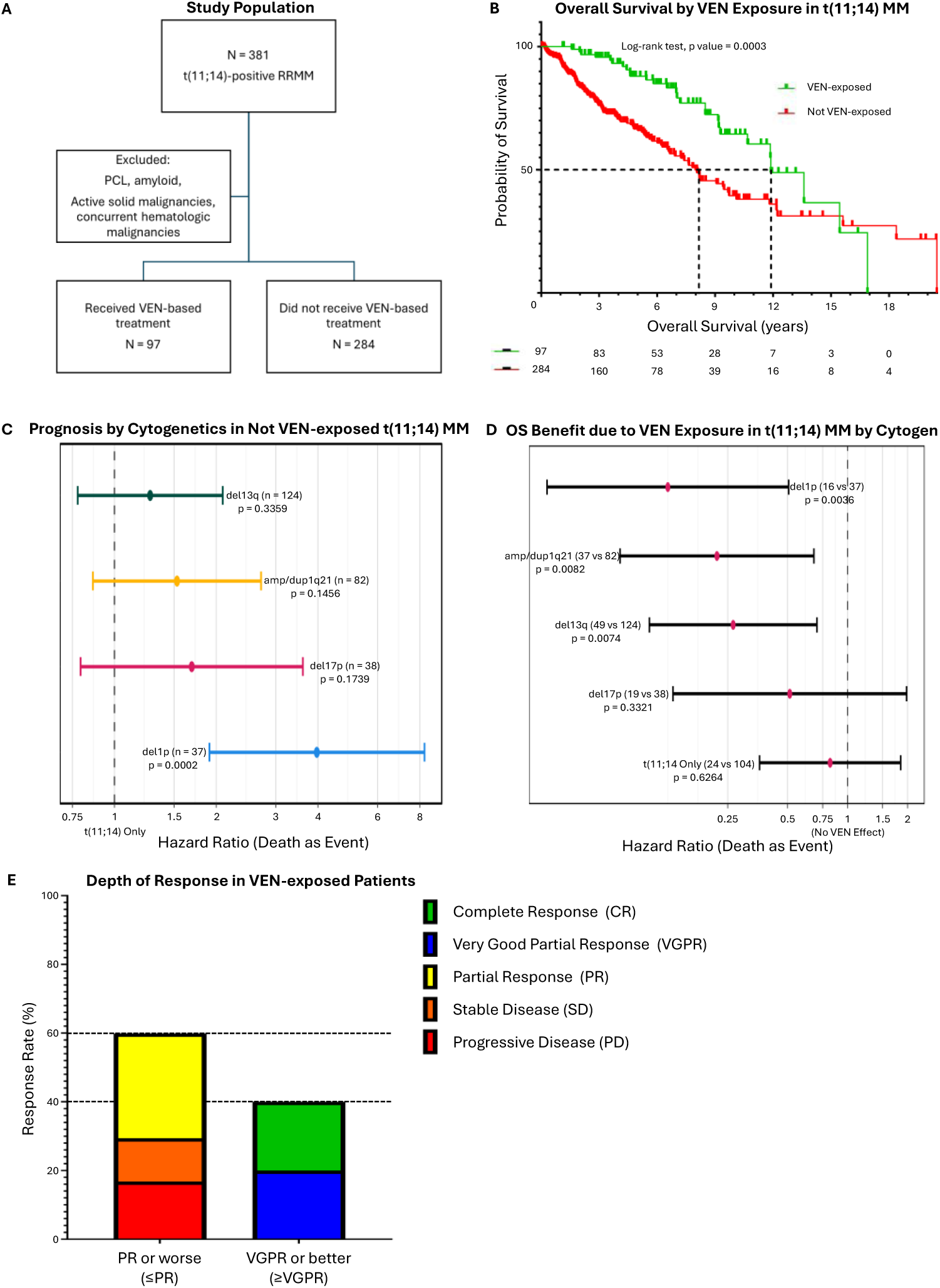
Exposure to VEN results in a consistent OS improvenent regardless of treatnent with drugs fron novel agent era/innunotherapies, ASCT, penta-exposure, and nunber of LOT. **(A)** An overview of the study population of MM patients with t(11;14), where patients with plasma cell leukemia, amyloid, and other concurrent cancers were excluded. **(B)** A Kaplan-Meier overall survival comparison between t(11;14) MM patients who received at least one VEN-based therapy for MM at any point following diagnosis and those who received other therapies. **(C)** A forest plot featuring multivariable Cox proportional hazards analyses by cytogenetics comparing OS of MM patients in the Not VEN-exposed group showing the prognosis of each additional cytogenetic abnormality in comparison with t(11;14) only (HR =1). **(D)** A forest plot featuring multivariable contrastive Cox proportional hazards analyses between VEN-exposed and Not VEN-exposed for each cytogenetic abnormality (HR = 1 is No VEN Effect). **(E)** A stacked bar plot showing the percent IMWG best response to a VEN-based therapy as PR or worse (≤PR) and VGPR or better (≥VGPR).

Using MM diagnosis date as time zero, OS was superior for VEN-exposed t(11;14) MM patients compared to the rest of the cohort in Kaplan-Meier analysis (log-rank test, p value = 0.0003; Fig. 1B). OS was also longer for VEN-exposed t(11;14) MM patients compared to Not-VEN-exposed MM patients within each of the subgroups that are over-represented in VEN-exposed patients (Supplementary Fig. 1). These data indicate that exposure to VEN results in consistent OS improvement regardless of treatment with novel agents, ASCT, penta-exposure, and the number of LOT. A recent study showed that the presence of cytogenetic abnormalities, amp/gain 1q and del17p, in t(11;14) MM patients can lead to worse outcomes (PFS) following VEN treatment^23^. This motivated us to explore the impact of concurrent cytogenetic abnormalities on OS in Not-VEN-exposed t(11;14) MM patients in our cohort. The results from a multivariable Fig. 1C. We noted significantly lower OS in Not-VEN-exposed t(11;14) MM patients with concurrent del1p (Cox Proportional Hazard – Cox-PH, p = 0.0002) compared to Not-VEN-exposed t(11;14) MM patients with no concurrent cytogenetic abnormalities (reference HR=1). Next, we determined the impact of VEN treatment on OS of t(11;14) MM patients in each cytogenetic abnormality subgroup using multivariable, contrastive CoxPH analyses (Not-VEN-exposed as reference HR=1) shown in Fig. 1D. Most notably, a significant improvement in OS was observed due to VEN treatment in del13q (Cox-PH, p = 0.0074), amp/gain1q21 (Cox-PH, p = 0.0082), and del1p (Cox-PH, p = 0.0036). These data suggest that exposure to VEN can improve OS in t(11;14) MM patients despite the poor prognosis conferred by concurrent cytogenetic abnormalities.

We also examined the efficacy of VEN-containing therapy in this relapsed MM cohort. VEN-based treatment resulted in a response in 69/95 patients (ORR 73%) and a very good partial response (VGPR) or better in 38/95 patients (40%, Fig. 1E). Deeper responses were also noted in 4/95 patients who achieved a stringent complete response or MRD-negative status. This response rate is slightly lower when compared to those reported in some clinical trials and real-world cohorts^14–26^.

Next, we evaluated how co-occurring cytogenetic abnormalities, expression of genes from the BCL2 family, and partner drugs affect the following endpoints in VEN-based regimens: PFS and PR or worse; (referred to as ≤PR). Consistent with the OS analysis, concurrent del1p was a major determinant of worse outcome (Cox-PH, p = 0.0074 for PFS, Supplementary Fig. 2A-B). Similarly, we assessed the impact of BCL2-family gene expression ratios (univariable CoxPH analyses, Supplementary Fig. 2C – D) and expression of BCL2-family of genes on VEN PFS and odds of ≤PR (multivariable CoxPH analyses for variables with Variance Inflation Factor ≤ 5, low multicollinearity, to only include mutually independent variables in the multivariable analysis; Supplementary Fig. 2E – F). The most significant relationships were noted between expression of PMAIP1 (NOXA, pro-apoptotic protein) and longer PFS (p = 0.0612) and between expression of MCL1 and shorter PFS (p = 0.0604). In this t(11;14) bearing cohort, BCL2 expression and its ratios with other BCL2-family genes yielded no associations for either PFS or ≤PR.

Collectively, our findings across a heterogeneous cohort of t(11;14) patients emphasize the importance of employing VEN earlier in disease course and the need to identify the critical factors linked to VEN response in t(11;14) MM patients. Additionally, our data also indicates VEN and DARA are optimal partners. This BCl2-inhibitor/anti-CD38 mAb combination was associated with significantly improved likelihood of a deeper response (p = 0.0022, lower odds ratio of ≤PR) relative to VEN+DEX only treated cohort and was non-significantly associated with longer PFS (p = 0.0912). In contrast, proteasome inhibitors and IMiDs had no significant associations with outcomes (Supplementary Fig. 2G – H).

### Treatnent-Associated Hypogannaglobulinenia Drives Infection Risk During VEN Therapy

Adverse events (AEs) related to VEN were observed in 33 of 97 patients (34%) and were predominantly hematologic (Fig. 1E). VEN was interrupted due to non-hematologic toxicity in 13 patients (14%) and permanently discontinued in 8 patients (8%). Infections occurred in 18 patients (19%), defined radiographically, microbiologically, or by strong clinical suspicion; most were bacterial (n=16), with one viral and one mixed bacterial/viral infection. Of these, seven patients required hospitalization for intravenous antibiotics, and one patient (1%) died of pneumonia while on VEN. Hypogammaglobulinemia (IgG < 500 mg/dL) was noted in 24 patients (25%) during treatment, yet only 4 patients (4%) received IVIG (intravenous immunoglobulin). Infections were significantly more common among patients with IgG < 500 compared to those with higher IgG levels (46% vs. 10%; p < .01), underscoring a potential role for prophylactic IVIG. The patient who died of pneumonia had IgG < 500 and was not receiving IVIG prophylaxis.

### Evolution of t(11;14) Multiple Myeloma Biology across Disease States by FISH Cytogenetics

We used clinical FISH results from 456 patients collected at different time points across disease states to investigate how genomic complexity impacted response to VEN-based therapy (N = 738). Based on the co-occurrence of secondary cytogenetic abnormalities with t(11;14), we classified 200 samples (with matched FISH and disease status) into t(11;14) simple (no secondary abnormalities, 30.77%), first-order complexity (one secondary abnormality, 41.02% – predominantly del13q and amp/gain1q21), second-order complexity (two secondary abnormalities, 19.49% – predominantly del13q-amp/gain1q21 and del13q-del17p), and third-order complexity (with three secondary abnormalities, 8.72% – predominantly del13q-amp/gain1q21-del17p). This is depicted in Fig. 2A as a stacked bar plot stratified by MM disease states – smoldering MM (SMOL), NDMM, early relapsed/refractory MM (ERMM), and late relapsed/refractory MM (LRMM). As shown in Fig. 2A, we demonstrated that the most frequent secondary cytogenetic abnormality was del13q (45.13%) closely followed by important to note that complexity, as defined here, is an abstraction to facilitate cohort-wide analyses. In Fig. 2B, we represent the change in composition of FISH complexity across disease states. This data illustrates an overall gain in complexity, as the percentage of t(11;14)-simple patients are significantly lower in LRMM compared to therapy-naïve patients and the converse is true for third-order cytogenetic complexity. Next, we characterized the incidence of FISH complexity using time since MM diagnosis for each sample (n = 738) using an inverted empirical cumulative distribution function (ECDF) in Fig. 2C. While it looks similar to a Kaplan-Meier survival plot, it depicts the time to incidence of FISH complexity and is not a measure of overall survival. For each sample, the time from initial MM diagnosis to the date of sample collection was used to assess when complexity was observed. This analysis revealed a statistically significant difference between complexity groups (log-rank test, p = 0.0016). Cases with simple cytogenetic profiles were observed predominantly early after diagnosis, whereas progressively higher levels of complexity tended to be detected later in the disease course.

**Fig. 2.**
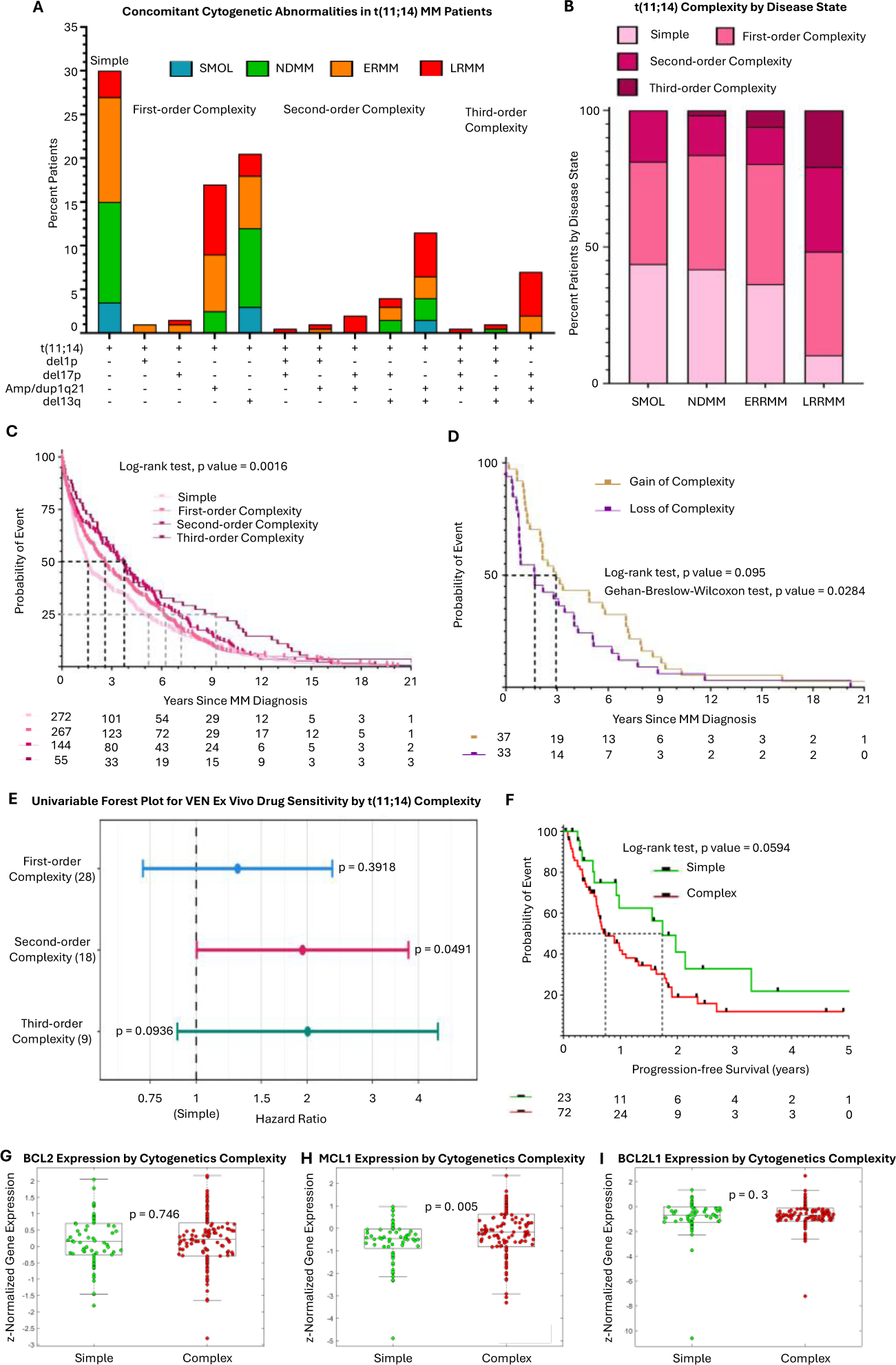
Evolution of t(11;14) MM and its role on VEN sensitivity. **(A)** A stacked bar plot showing percent patients in each disease status across concomitant cytogenetic abnormalities in t(11;14) MM patients, where the number of concomitant cytogenetic abnormalities was used to classify t(11;14) MM patients into groups of complexity. **(B)** A stacked bar plot showing percent patients in each t(11;14) complexity group by disease status. **(C)** A Kaplan-Meier plot comparing time to complexity event since MM diagnosis. **(D)** A Kaplan-Meier plot comparing time to gain or loss of complexity event from MM diagnosis. **(E)** A multivariable forest plot showing the association between VEN ex vivo sensitivity and order of complexity in reference to the Simple group (HR =1). **(F)** A Kaplan-Meier plot comparing PFS of VEN treated MM patients between simple and complex t(11;14) groups. **(G)** – **(I)** Box plots comparing BCL2, MCL1, and BCL-XL expression in t(11;14) MM patients between simple and complex groups.

To investigate how complexity changes over time within individual patients, we examined sequential FISH biopsies from 200 MM patients. We identified three patterns: gain of complexity (n = 37, 18.5%), loss of complexity (n = 33, 16.5%), and no change (n = 130, 65%). Although the majority of patients showed stable cytogenetic profiles, those who experienced a change were approximately equally likely to gain or lose complexity. Notably, loss of complexity tended to occur earlier than gain, although this difference did not reach statistical significance (log-rank test, p = 0.095) (Fig. 2D).

We further examined the relationship between FISH-defined cytogenetic abnormalities and ex vivo VEN sensitivity (see *Ex Vivo Drug Sensitivity Assays* in Methodology) in all patients with paired data (N = 362, cytogenetically unselected cohort, Cox-PH analyses shown in Supplementary Fig. 3A). The presence of t(11;14) was significantly associated with increased sensitivity (lower AUC) to VEN (HR = 0.5), whereas t(14;16), t(4;14), del13q, and amp/gain 1q21 were linked to resistance. Consistent with findings above, del13q and amp/gain 1q21 are the most frequent secondary abnormalities co-occurring with t(11;14), and ex vivo these abnormalities contribute to VEN resistance. Next, we examined the relationship between cytogenetic complexity and ex vivo VEN response in 76 t(11;14) MM samples (cytogenetically selected sub-cohort from the N = 362 cohort described above). We observed a significant association with VEN resistance in second-order complexity (p = 0.0491) and a non-significant yet notable trend for third-order complexity (p = 0.0936) compared to t(11;14)-simple (Fig. 2E). Notably, cytogenetic complexity was also associated with clinical resistance (in addition to ex vivo resistance) to VEN. In exploring the clinical cohort of t(11;14) MM patients (N = 95), we found that t(11;14)-simple MM patients had a statistically non-significant, but clinically meaningful improvement in median PFS of nearly one year compared to those with complex FISH profiles (log-rank test, p = 0.0594, Fig. 2F).

Finally, we designed a regression tree model, which was trained using cytogenetic data from FISH as input and *ex vivo* VEN response as output (Supplementary Fig. 4) to classify the same 95 VEN-exposed t(11;14) MM as either VEN sensitive or VEN resistant (see *Predictive machine learning models* in Methods). The model classification (Supplementary Fig. 3B) was identical to FISH classification into t(11;14)-simple and t(11;14)-complex (Fig. 2F). Complexity by FISH, as opposed to a specific cytogenetic abnormality, emerged as a predictor of ex vivo VEN resistance from unsupervised training of the regression tree model suggesting that the association with complexity is likely stemming from evolutionary selective advantage conferred by concomitant cytogenetic abnormalities driving poor prognosis and not the emergence of a target-specific resistance mechanism driven by a singular cytogenetic abnormality. Consistent with this inference, no significant differences were observed in BCL2 and BCL2L1 expression between simple and complex patients, while complex patients have a higher MCL1 expression (also seen in Supplementary Fig. 2E).

### CD1/CD2 Exclusivity Improves Outcomes in Venetoclax-treated Multiple Myeloma Patients

In our prior work, we developed an unsupervised approach to identify transcriptional programs associated with ex vivo drug response in MM^28^. We employed this approach here with a particular focus on VEN, in Supplementary Fig. 3C, we present a waterfall plot depicting the gene-wise correlation between expression levels and ex vivo response to VEN in a cohort of 127 cytogenetically unselected MM patients with paired RNA-seq and ex vivo sensitivity data. Consistent with known biology, CCND1 and BCL2 expression were strongly correlated with ex vivo sensitivity to VEN, whereas BCL2L1 and MCL1 expression were highly correlated with resistance. To further delineate coordinated gene expression programs underlying these responses, we conducted GSEA, which identified clusters of co-expressed genes associated with ex vivo resistance or sensitivity (depicted as red and blue clusters, respectively, in Supplementary Fig. 3D). Using the methodology outlined in Supplementary Fig. 4 (see *Predictive machine learning models* in Methodology), we trained a regression tree model based on the median expression of these gene clusters and corresponding ex vivo responses to classify patients as predicted sensitive or predicted resistant to VEN. In Supplementary Fig. 3E, we compared PFS among VEN-treated patients stratified by this model, observing a statistically significant improvement in PFS for patients predicted to be sensitive compared with those predicted to be resistant (p = 0.0001, log-rank test). Importantly, as shown in Supplementary Fig. 3F, patients predicted to be resistant exhibited significantly higher ssGSEA (single-sample gene set enrichment analysis) enrichment scores for the LB program (MM subtype characterized by three or more bone lesions, identified using microarray data by a study from University of Arkansas Medical Sciences, UAMS^27^) compared with predicted-sensitive patients (p = 0.0004, unpaired t-test).

The UAMS classification system^27^ relies on gene expression profiling to allocate MM tumors into one of seven subgroups, where five of them were associated with primary cytogenetic abnormalities – CD1/CD2 with t(11;14), MS with t(4;14), MF with t(14;16), and HD with hyperdiploidy; and the remaining two were functional (non-genomic) programs– PR represents proliferation and LB represents greater than three bone lesions^27^. These subgroups have been demonstrated to have prognostic value in NDMM patients with PR, MF, and MS subtypes associated with poor prognosis^27,29^, and more recently LB in African Americans^27,29,30^. We have classified 24 pre-VEN-treatment RNAseq samples (only 24 of the 97 VEN-exposed patients had RNAseq done before start of treatment) from the (11;14) MM clinical cohort (also used in Supplementary Fig. 2C – F) using single-sample gene set enrichment analysis (ssGSEA) calculated on UAMS gene sets. In Fig. 3A, we present a heatmap of ssGSEA enrichment scores for each subtype (in rows) and patients (in columns) with bar plots below, representing the patients’ cytogenetic abnormality status. We observe three distinct subgroups of t(11;14) MM patients – CD1/CD2-exclusive (positive enrichment for CD1 & CD2 and no/negative enrichment for other groups), CD1/CD2 + MF/PR (positive enrichment for CD1 & CD2 along with positive enrichment for either MF/PR and no/negative enrichment for LB), and CD1/CD2 + LB (positive enrichment for CD1 & CD2 along with positive enrichment for LB, regardless of enrichment for any other group). From a transcriptomic standpoint, CD1/CD2-exclusive represents a form of functionally simpler biology, and CD1/CD2 + MF/PR and CD1/CD2 + LB represent a form of functional complexity, as the latter do not imply a loss of CD1/CD2 programs, but addition of MF/PR or LB phenotypes.

**Fig. 3.**
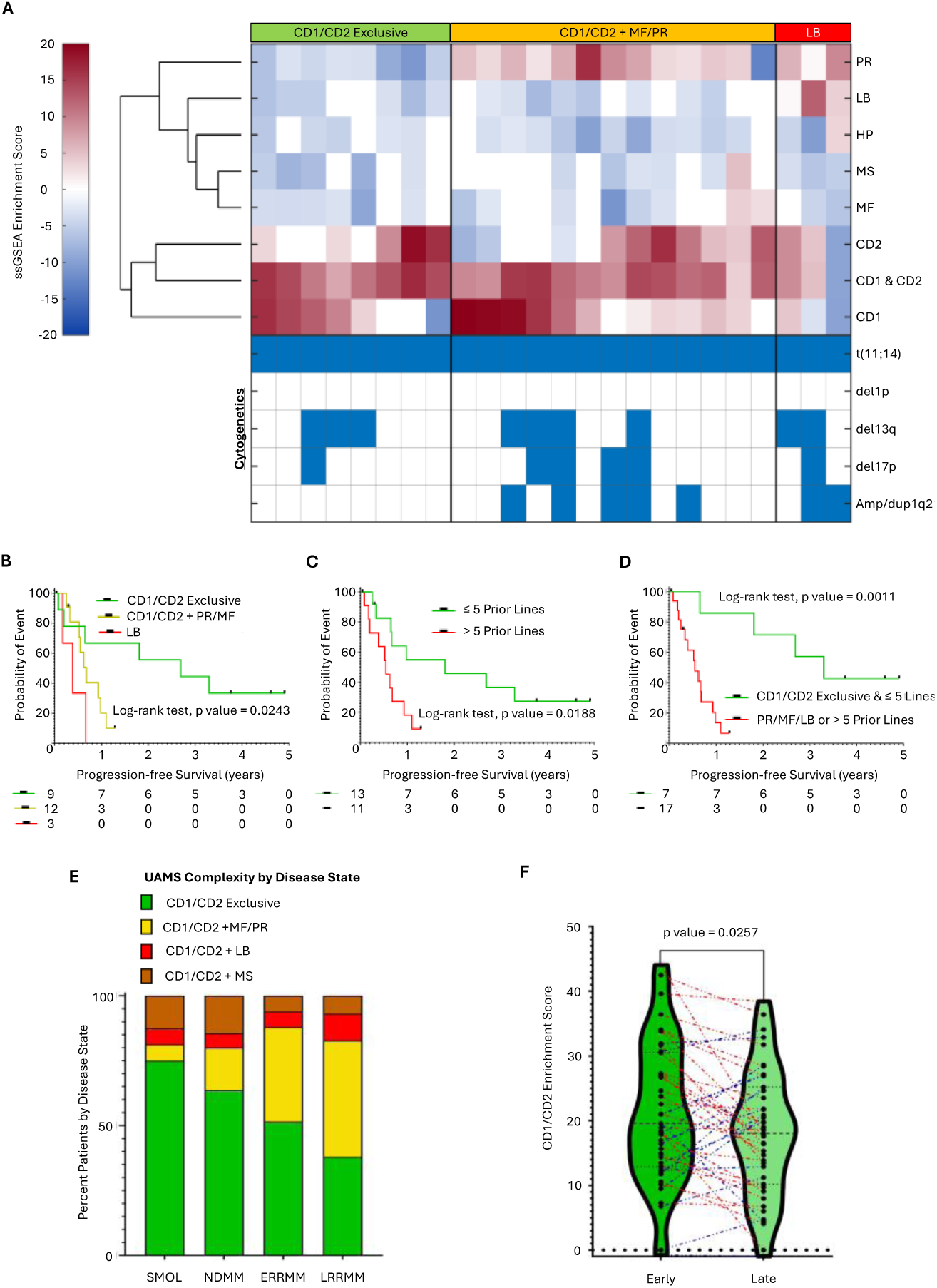
Evolution of functional conplexity in t(11;14) MM and bionarkers for VEN response. **(A)** Clustergram showing ssGSEA enrichment scores for each gene set in rows and patients in columns. **(B)** Kaplan-Meier survival comparison of PFS between CD1/CD2 exclusive, CD1/CD2 + MF/PR, and CD1/CD2+LB. **(C)** Kaplan-Meier survival comparison of PFS between MM patients with less than or equal to 5 and greater than 5 prior lines of therapy. **(D)** Kaplan-Meier PFS comparison between CD1/CD2 exclusive and <=5 prior LOT and the remaining cohort. **(E)** Stacked bar plot of percent patients with CD1/CD2 Exclusive and functionally complex groups for each disease state. **(F)** Violin plot of CD1 & CD2 enrichment scores between two sequential biopsies using a paired t-test.

When we explored the outcomes of these three molecularly-defined subtypes (Fig. 3B), the CD1/CD2-exclusive group exhibited significantly longer PFS with VEN-based therapy (log-rank test, p = 0.0243, median 2.69 years, compared to median 0.67 years for CD1/CD2 + MF/PR and median 0.38 years for CD1/CD2 + LB). Additionally, we observed differences in prior treatment exposure at the time of VEN initiation: the CD1/CD2-exclusive group was the least heavily pretreated (median 4 prior LOT), followed by the CD1/CD2 + MF/PR group (median 6 prior LOT), and the CD1/CD2 + LB group (median 7 prior LOT). This observation prompted us to explore the association between PFS and prior LOT (Fig. 3C). Using a cut-off of less than or equal to five prior LOT (owing to its prognostic impact shown in Supplementary Fig. 1A – B), we were able to identify a subset of t(11;14) MM patients with significantly longer PFS on VEN (log-rank test, p = 0.0188, median 1.8 years, compared to the group with >5 prior LOT, median 0.54 years) regardless of transcriptomic profile. We identified the highest discriminatory value using a composite criterion and we found that CD1/CD2-exclusive t(11;14) MM patients with ≤ 5 prior LOT had the longest PFS (log-rank test, p = 0.0011) compared to others (Fig. 3D). Next, we studied the incidence rates of CD1/CD2 exclusive, CD1/CD2+MF/PR, and CD1/CD2+LB in a larger t(11;14) MM cohort (N = 195, with paired FISH and RNAseq drawn from the larger Moffitt cohort). In addition to the three molecular subtypes mentioned above, we found a fourth low-frequency subtype, CD1/CD2+MS (no/negative enrichment for other subtypes), which we have included in subsequent analyses. In Fig. 3E, we show a stacked bar plot illustrating that the proportion of CD1/CD2-exclusive patients progressively decreases across disease stages — from SMM to NDMM to ERMM to LRMM — while the proportion of PR-enriched patients increases correspondingly. Finally, in Fig. 3F, we demonstrate the reduction of CD1/CD2-exclusivity in a cohort of 43 t(11;14) MM patients (drawn from the 195-cohort with paired FISH and RNAseq data) with sequential RNAseq samples, we observed a statistically significant decrease in CD1 & CD2 enrichment scores between early and late biopsies (paired t-test, p = 0.0257).

### Characterizing the resistance phenotypes in t(11;14) sanples

We characterized the biology of 174 t(11;14) active MM tumors (NDMM, ERMM, and LRMM; drawn from the 195-cohort with paired FISH and RNAseq), which were classified as CD1/CD2 exclusive (N = 83) or CD1/CD2+PR/MF/MS/LB (N = 91). Agglomerative clustering of z-normalized ssGSEA scores of Cancer Hallmarks was conducted for samples in each of these molecular subtypes, which yielded two distinct clusters in each molecular subtype (Fig. 4A), Group 1 and Group 2 in CD1/CD2 exclusive and Group 3 and Group 4 in CD1/CD2+PR/MF/MS/LB. Based on the cancer hallmarks enriched in each group, similar biology can be noted between Group 1 and Group 3 (immune/microenvironment dependency), and Group 2 and Group 4 (cell cycle, MYC hyperactivation, and proliferation). In our prior work^31^, we identified these two mechanisms to be associated with disease progression in MM, i.e. distinct survival mechanisms MM cells can adopt. Despite the similarities between Group 1 and Group 3, we note that Group 3 has higher enrichment scores (and lower VEN PFS in the clinical cohort, p = 0.0483, Supplementary Fig. 5) for the immune/microenvironment-dependent biology compared to Group 1. Similarly, we note that Group 4 has higher enrichment scores for the cell cycle/proliferation biology compared to Group 2. Notably, despite the biological differences between Groups 1 and 2, and 3 and 4; no significant differences in VEN PFS were observed in the clinical cohort (Supplementary Fig. 5). Disease status and cytogenetic abnormalities are shown for each sample below the heatmaps featuring UAMS z-normalized ssGSEA scores and z-normalized expression of CCND1, BCL2, BCL2L1, and MCL1 genes in Fig. 4A. No significant associations between each of the four groups and a specific cytogenetic abnormality or disease status can be noted, while Group 4 has higher incidence of LRMM patients and higher-order complexity by FISH. Violin plots comparing z-normalized ssGSEA scores for UAMS gene sets are shown in Fig. 4B – G, where CD2 is significantly associated with Groups 1 and 3 (immune/microenvironment dependence), MF and MS are enriched in Group 3, and PR is selectively enriched in Group 4 (despite sharing similar MYC activation biology with Group 2). Violin plots (Fig. 4H – N) comparing z-normalized gene expression between the four groups show a significant decrease in BCL2 expression (p = 0.0018) and a non-significant yet notable decrease in BCL2L11 (BIM) expression (p = 0.0975) in Group 4 compared to Group 2. This may contribute to inferior outcomes to VEN treatment in the Group 4 subset of functionally complex t(11;14) MM patients with the loss of target expression BCL2 and a downstream activator protein BIM (activates BAX/BAK to engage in mitochondrial outer membrane permeabilization). A second pattern of resistance was seen in Group 3 compared to Group 1. Here, we observe a significant decrease in CCND1 expression (p = 0.0006), a non-significant yet notable increase in BCL2A1 expression (p = 0.0553), and a significant increase in HIF1A (p = 0.0006), which suggests an enrichment for hypoxia-driven EMDR stress adaptation in Group 3 compared to Group 1 and BCL2A1 also sequesters BIM (similar to BCL2) dampening the downstream effect of BCL2-inhibition.

**Fig. 4.**
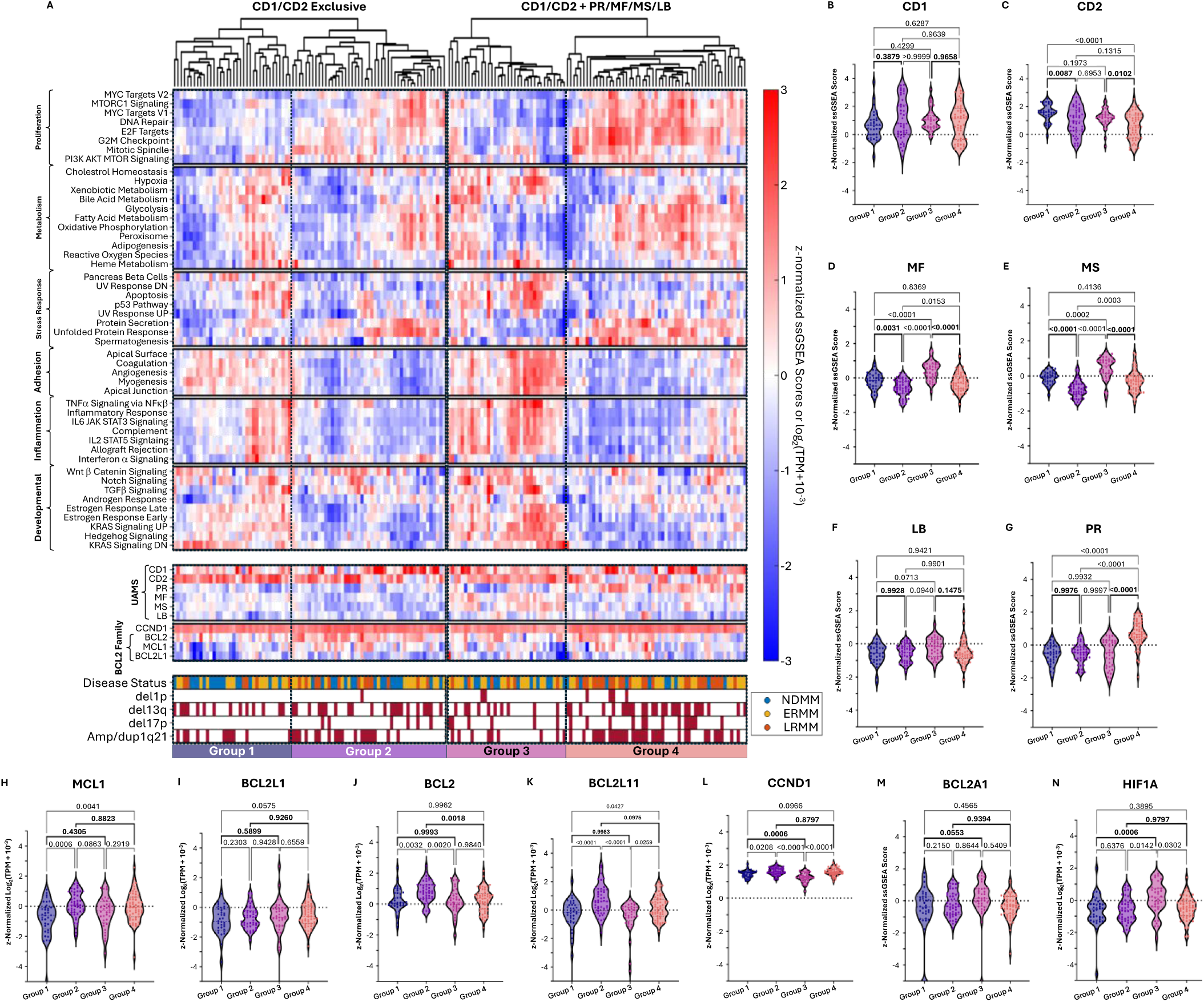
Characterization of the CD1/CD2 Exclusive and CD1/CD2¦PR/MF/MS/LB groups. **(A)** A bipartite clustergram of z-normalized ssGSEA scores (top) of Cancer Hallmarks for CD1/CD2 Exclusive and CD1/CD2+PR/MF/MS/LB independently. The clustering identified two sub-groups in each group, resulting in Group 1 and Group 2 under CD1/CD2 Exclusive, and Group 3 and Group 4 under CD1/CD2+PR/MF/MS/LB. Heatmaps (below) of UAMS z-normalized ssGSEA scores and z-normalized log_2_(TPM+10^-3^) expression of CCND1, BCL2, MCL1, and BCL2L1 genes. Disease status and cytogenetic abnormality status were added as bar plots below the heatmaps. **(B)** – **(G)** Violin plots comparing z-normalized ssGSEA scores between Groups 1 – 4 of UAMS gene sets CD1, CD2, MF, MS, LB, and PR; respectively. **(H) – (N)** Violin plots comparing z-normalized log_2_(TPM+10^-3^) expression between Groups 1 – 4 of MCL1, BCL2L1, BCL2, BCL2L11, CCND1, BCL2A1, and HIF1A genes; respectively.

### Novel Therapies Targeting Functionally Conplex t(11;14) Multiple Myeloma Biology

Although CD1/CD2-exclusive t(11;14) patients show encouraging responses to VEN, the prognosis of other molecular subtypes remains poor. To identify potential therapeutic vulnerabilities in these groups, we leveraged paired RNAseq and ex vivo response data for 76 t(11;14) MM samples to identify standard-of-care (SOC) single agents and combinations, and experimental agents that are selectively efficacious in functionally complex t(11;14) subtypes. For this analysis, we expanded our mutually exclusive three-group classification from Fig. 3A with non-mutually exclusive molecular subtypes: CD1/CD2-exclusive, CD1/CD2+PR/MF/MS/LB, CD1/CD2+PR, CD1/CD2+LB, CD1/CD2+MF, CD1/CD2+MS, Group 3, and Group 4 to identify drugs that show selective sensitivity in a subtype-specific manner. In Fig. 5A, we present a heatmap of mean z-normalized ex vivo responses to a panel of therapies and combinations (normalization was done per drug across all MM samples tested with that drug ex vivo, while the heatmap shows data for only t(11;14) cohort; ex vivo responses were quantified as area under the curve, AUC, see *Ex Vivo Drug Sensitivity Assays* in Methodology). Comparisons of AUC z-scores between CD1/CD2-exclusive group and each of the functionally complex subtypes were carried out for each SOC and experimental agents. Complex subtypes with a statistically significant difference (at α = 0.1) in z-normalized AUCs (relative ex vivo sensitivity) are marked with asterisks. Compared to the CD1/CD2-exclusive group, the CD1/CD2+PR/MF/MS/LB group shows increased sensitivity to IXA, DEFA (FAK and PYK2), THZ1 (CDK7), DARA+LEN, and IXA+LEN, and resistance to VEN and CFZ+LEN (Supplementary Fig. 6 shows box plots with individual data points).

**Fig. 5.**
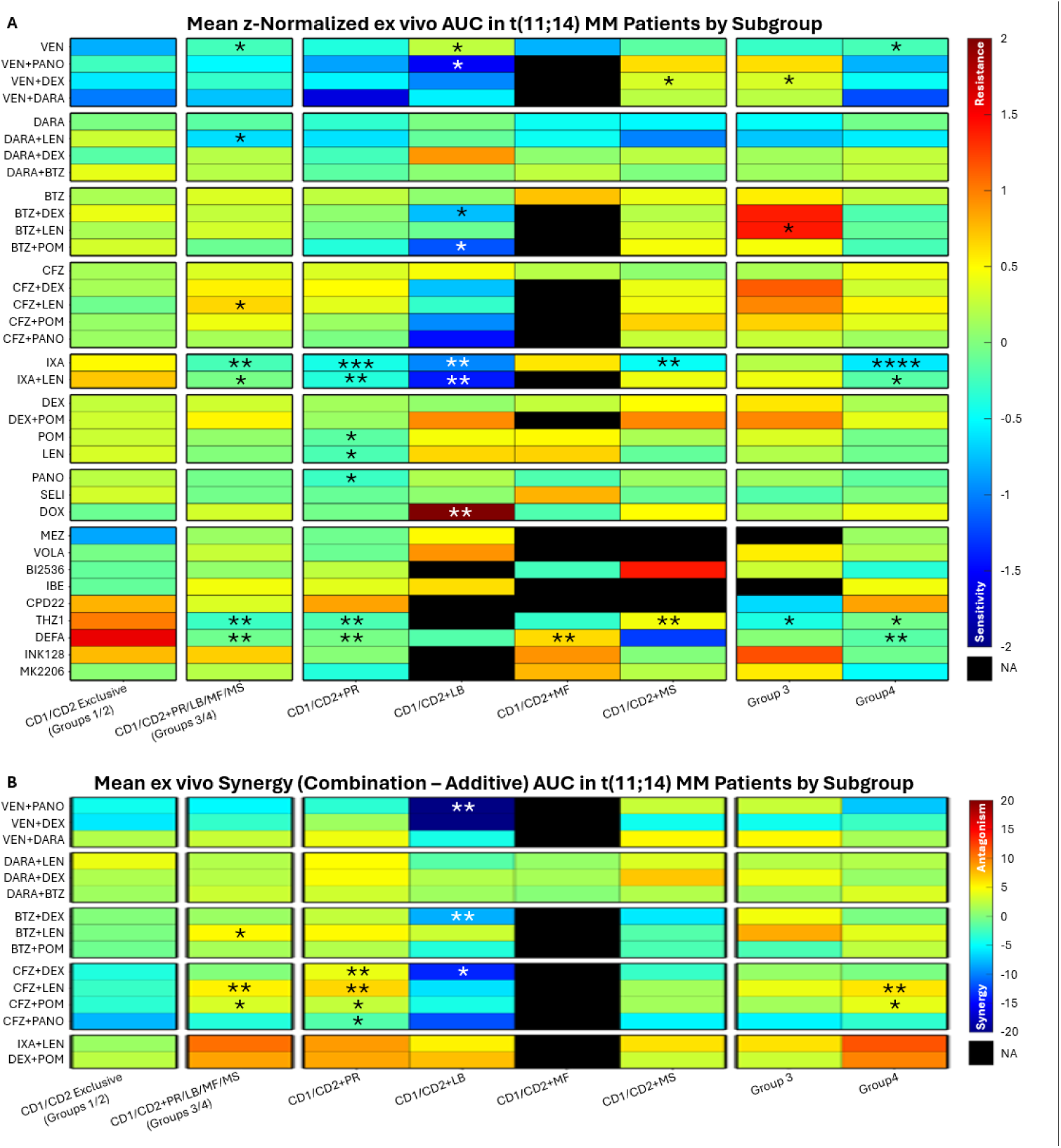
Transcriptonic subtype-specific therapy selection in t(11;14) MM inforned by ex vivo drug sensitivity assays. **(A)** Heatmap of mean z-normalized AUC for SOC and experimental agents, and their combinations in 76 patients with paired RNAseq and ex vivo response. **(B)** Heatmap of mean synergy (combination AUC – additive AUC). * signifies p ≤ 0.1, ** signifies p ≤ 0.05, *** signifies p ≤ 0.005, and **** signifies p ≤ 0.0005.

In Fig. 5B, we show a heatmap of mean synergy (defined as combination AUC minus additive AUC, estimated using Bliss statistical independence^32^) across two-drug combinations tested ex vivo. We observed statistically significant synergy in the CD1/CD2 + LB group for BTZ+DEX combinations (Supplementary Fig. 7 shows box plots with individual data points).

## Discussion

In this large real-world cohort of patients with t(11;14) MM, exposure to VEN was associated with a clinically meaningful improvement in OS (Fig. 1B). This association was observed despite enrichment of the VEN-exposed group for higher number of prior LOT and selective exposure to contemporary agents and immunotherapies (Table 1 and Supplementary Fig. 1). VEN exposure was significantly associated with longer OS in patients harboring secondary high-risk cytogenetic abnormalities (del1p, amp/gain1q21, and del13q; otherwise associated with poor OS), suggesting that VEN exposure confers durable clinical benefit across a range of disease severities within the t(11;14) subgroup (Fig. 1C – D). VEN treatment was associated with predictable toxicity, chiefly infectious complications. Infections occurred in a minority of patients and were more frequent among those with hypogammaglobulinemia during therapy, particularly when IgG levels fell below 500 mg/dL. Intravenous immunoglobulin prophylaxis was infrequently utilized in this cohort. These findings suggest that infection risk during VEN therapy may reflect underlying immune vulnerability and cumulative treatment exposure and may be mitigated through closer immunologic monitoring and supportive care, although this warrants prospective evaluation. These observations suggest that t(11;14) MM patients exposed to VEN at any point following diagnosis experience better prognosis across a range of disease severities in t(11;14) MM, while notably the positive influence is enhanced with the introduction of VEN in earlier lines of therapy. This variability prompted further examination of underlying biological features, specifically cytogenetic and transcriptional states that may influence the persistence of VEN PFS over time.

t(11;14) multiple myeloma is recognized as an intermediate-risk entity in the era of IMiDs, proteasome inhibitors, and monoclonal antibodies. However, the impact of concomitant cytogenetic abnormalities on outcomes to VEN treatment within t(11;14) MM remains incompletely characterized. We conducted longitudinal cytogenetic analyses to show that secondary cytogenetic abnormalities accumulate with disease progression (Fig. 2A – B), resulting in increasing genomic complexity that showed a significant association with ex vivo VEN resistance and a non-significant trend towards shorter PFS (Fig. 2E – F), where the low frequency cytogenetic abnormality del1p was significantly associated with shorter OS (Fig. 1C) and PFS (Supplementary Fig. 2A). These data suggest that the co-occurrence of del1p in t(11;14) MM leads to a relatively high-risk biology.

As a next step, we performed transcriptomic profiling, which revealed substantial functional heterogeneity within the t(11;14) MM population. Tumors segregated into historical UAMS CD1/CD2-exclusive states and functionally complex states characterized by the additive emergence of distinct transcriptional programs (PR/MF/MF/LB). This divergence provides a potential explanation for heterogeneity in VEN response and durability that is not captured by cytogenetics, individual gene expression, or BCL2 family ratios alone. It is clear that CD1/CD2-exclusive tumors demonstrating more durable responses compared with transcriptionally complex subgroups (Fig. 3B), which tend to occur at higher frequencies in later LOT. From a clinical standpoint, line of therapy may serve as a pragmatic surrogate for functional state when transcriptomic profiling is unavailable. In our cohort, earlier lines of therapy were enriched for CD1/CD2-exclusive biology and more durable VEN sensitivity, whereas later lines (>5) were increasingly characterized by transcriptional complexity and diminished durability VEN-based treatment (Fig. 3C – D). Furthermore, paired RNA-seq analyses showed progressive loss of CD1/CD2 exclusivity with advancing disease and increasing prior LOT (Fig. 3E – F).

Further characterization of these transcriptional states showed that CD1/CD2-exclusive and CD1/CD2+PR/MF/MS/LB tumors each partitioned into two convergent biologically-defined programs: an immune/microenvironment-dependent program (Group 1 and Group 3) and a cell-cycle/proliferation program (Group 2 and Group 4, Fig. 4A). The functionally complex counterparts (Group 3 and Group 4) showed stronger enrichment for their respective programs that were found to be survival mechanisms adopted by MM cells during disease progression^31^. Group 3, the functionally complex, immune/microenvironment dependent molecular subtype resembles an environment-mediated drug resistant (EMDR) state^33^, which was marked by MF/MS enrichment (Fig. 4D – E), decreased CCND1 expression (Fig. 4L), increased HIF1A expression (Fig. 4N), and a trend toward increased BCL2A1 expression (Fig. 4M). These findings support a hypoxia-associated stress-adaptation state in which alternative BIM sequestration may blunt the downstream effect of BCL2 inhibition. Group 4, the functionally complex cell cycle/MYC-activation/proliferation program was simultaneously enriched for glycolysis, oxidative phosphorylation, fatty acid metabolism, and other metabolic pathways; suggesting a bioenergetic state, where the cell taps multiple sources of energy to support rapid cell proliferation. Group 4 was selectively enriched for PR biology (Fig. 4G) and showed significantly decreased BCL2 expression (Fig. 4J) with a trend toward decreased BCL2L11 (BIM) expression (Fig. 4K) compared with its CD1/CD2-exclusive counterpart, Group 2. These findings provide a mechanistic basis for reduced VEN durability in transcriptionally complex disease, linking inferior response to both loss of BCL2/BIM-dependent apoptotic priming and acquisition of stress-adaptive or proliferative/hypermetabolic survival programs.

Ex vivo drug sensitivity profiling provided additional functional context for these transcriptomic states. Sensitivity patterns varied by functional subtype, with CD1/CD2-exclusive samples demonstrating greater Ven sensitivity and functionally complex states exhibiting relative resistance and alternative vulnerabilities (Fig. 5A). These ex vivo findings were concordant with observed differences in clinical durability and support the interpretation that transcriptional state captures functional dependencies not reflected by cytogenetics alone. Our results suggest that no single VEN partner is likely to be uniformly effective across patients or disease states. Ex vivo drug sensitivity profiling identified state-specific vulnerabilities that varied by functional subtype. Most notably, the selective sensitivity of IXA (not BTZ/CFZ) for the PR-enriched Group 4 was striking, which can be attributed to a distinct subgroup-specific reliance on the β1/β1i proteasomal subunits^34^ (as opposed to β5/β5i specific targeting by BTZ/CFZ). In the clinical cohort, and consistent with previous studies highlighting complementary activity between agents^23^, we noted an association between DARA exposure with VEN+DEX and longer PFS (Supplementary Fig. 2G). Accordingly, in a molecularly unselected cohort DARA (or other anti-CD38 mAb) may be an optimal partner drug for VEN in CD1/CD2 exclusive t(11;14) MM. In case of PR-driven or LB-driven complexity, the combination DARA/VEN/IXA/DEX may be considered. Perhaps, the most difficult to treat subgroup may be Group 3 being resistant to nearly all agents/combinations tested ex vivo, except THZ1, which warrants preclinical studies confirming selective sensitivity in Group 3 to epigenetic modulators. These findings are hypothesis-generating and intended to inform prospective, biomarker-driven study design rather than immediate clinical decision-making.

Taken together, these results suggest that t(11;14) MM patients VEN-based therapy should be integrated at some point in their disease course. More specifically, VEN-based regimens must be considered sooner than 5 prior LOT. In the future, where RNAseq-based molecular subtyping can confirm the tumor state facilitating biomarker-driven therapy in t(11;14) MM more personized integration can be imagined. Prospective studies should therefore prioritize biological state, disease evolution, and treatment sequencing rather than relying solely on binary cytogenetic inclusion.

While well founded and hypothesis generating, several limitations should be considered when interpreting these findings. First, the retrospective design of this study introduces the potential for residual confounding, including treatment selection bias and time-dependent effects that cannot be fully accounted for despite adjustment for known clinical variables. Second, while this represents the largest real-world cohort of t(11;14) MM examined to date, subset analyses, particularly those involving specific cytogenetic abnormalities, transcriptomic subtypes, and ex vivo drug sensitivity profiles, were limited by sample size and should be interpreted as hypothesis-generating. Third, transcriptomic and functional analyses were performed on selected samples and may not capture the full spectrum of intra-patient heterogeneity or temporal evolution within the same tumor. In addition, ex vivo drug sensitivity does not necessarily recapitulate in vivo pharmacodynamics, microenvironmental interactions, or treatment tolerability. Finally, although our integrative framework offers a biologically coherent interpretation of VEN response heterogeneity, it remains speculative and requires prospective validation in biomarker-driven clinical trials designed to assess optimal timing, sequencing, and combination strategies.

## Acknowledgenents

The authors sincerely thank our multiple myeloma patients and their families for donating their samples for research purposes. We also thank the members of the Shain, Silva and Cleveland labs, and the Pentecost Family Myeloma Research Center. This research was made possible through the Oncology Research Information Exchange Network (ORIEN) Avatar Project in collaboration with Aster Insights (formerly known as M2Gen), the Total Cancer Care protocol and supported by the Tissue Core, Molecular Genomics Core, Cancer Pharmacokinetics and Pharmacodynamics Core, Biostatistics and Bioinformatics Core and Collaborative Data Services Core shared resource facilities at the H. Lee Moffitt Cancer Center & Research Institute; an NCI designated Comprehensive Cancer Center (P30-CA076292). This work was supported by philanthropic funding from the Pentecost Family Myeloma Research Center (to R.B., K.S., A.S.), and in parts by the H. Lee Moffitt Cancer Center Physical Sciences in Oncology (PSOC) Grant 1U54CA193489-01A1 (to K.S., A.S.), H. Lee Moffitt Cancer Center’s Team Science Grant (A.S., K.S.), Miles for Moffitt Foundation (A.S.), 2021 Multiple Myeloma Research Foundation (MMRF) Research Fellowship Award (P.S.), Florida Department of Health, Public Health Research, Biomedical Research Program (K.S.), and the Cancer Center Support Grant P30-CA076292 to the Moffitt Cancer Center. The funding agencies have not played any role in study design, data collection, data analysis, interpretation, writing of the report, and the decision to submit it for publication.

## Author Contributions

P.S., F.I., R.B., K.S., A.S., and A.G. contributed to the conception of the manuscript; F.I., D.D., R.I., S.T., P.P. prepared clinical metadata from clinical notes for the 381-patient t(11;14) MM cohort; R.R., M.S., M.M., X.Z., and A.P. conducted ex vivo assays; P.S. and A.S. conducted transcriptomic analyses and analyzed ex vivo data from the assays; P.S. and F.I. compiled data, carried out analyses, and wrote the manuscript; R.B., K.S., A.S., and A.G. provided mentorship and guidance. All authors have read, reviewed, and approved the manuscript.

## Data availability

The raw FASTQ files of RNAseq data used in this article are deposited in dbGAP under the accession number **phs003892** and supplementary data files are provided with all the clinical and ex vivo metadata required to reproduce the results.

## Code availability

The scripts for all codes used to generate the plots in this manuscript are attached as Supplementary Data with this manuscript along with the data files to reproduce all figures and supplementary figures. Scripts are in R (for survival analysis forest plots) and MATLAB (for heatmaps of transcriptomic data), and GraphPad Prism (for Kaplan-Meier and bar plots).

## Declarations

### Ethics approval and consent

Patient samples were used in accordance with the Declaration of Helsinki, International Ethical Guidelines for Biomedical Research Involving Human Subjects (CIOMS), Belmont Report, and U.S. Common Rule.

### Consent for publication

Not applicable.

### Conpeting Interests

R.B. reports research funding to the institution from Janssen, AbbVie, Bristol Myers Squibb, Regeneron, and Cell Centric, outside the submitted work; Also, member of advisory board for Janssen and Bristol Myers Squibb. K. S. reports honoraria from Amgen, AbbVie, Adaptive, Bristol Myers Squibb, GSK, Janssen, Regeneron, Sanofi, Sebia, and Takeda, and research funding to the institution from AbbVie, Pfizer and Karyopharm Therapeutics, not related to the submitted work. A.G. reports honoraria from Amgen, Janssen, Sanofi, Bristol Myers Squibb, Regeneron, Glaxo Smith Kline, Cellectar, and Pfizer and research funding to the institution from Janssen, outside of the submitted work. The other authors declare no competing interests.

