## Supplementary Figures for "Venetoclax-based Therapy Improves Outcomes across the Evolving Biology of t(11;14) Multiple Myeloma"

**Supplementary Fig. 1 |** Kaplan-Meier overall survival comparisons between t(11;14) MM patients who received VEN-based therapies and those who received other therapies for subgroup of patients that had statistically significant differences in proportions between the two groups as shown in Table 1, namely among patients who have **(A)** less than or equal to five prior lines of therapy, **(B)** greater than five prior lines of therapy, **(C)** exposed to Bortezomib, **(D)** exposed to Carfilzomib, **(E)** exposed to Lenalidomide, **(F)** exposed to Pomalidomide, **(G)** exposed to Daratumumab, **(H)** received CAR-T cell therapy, **(I)** received ASCT, and **(J)** were penta-exposed.

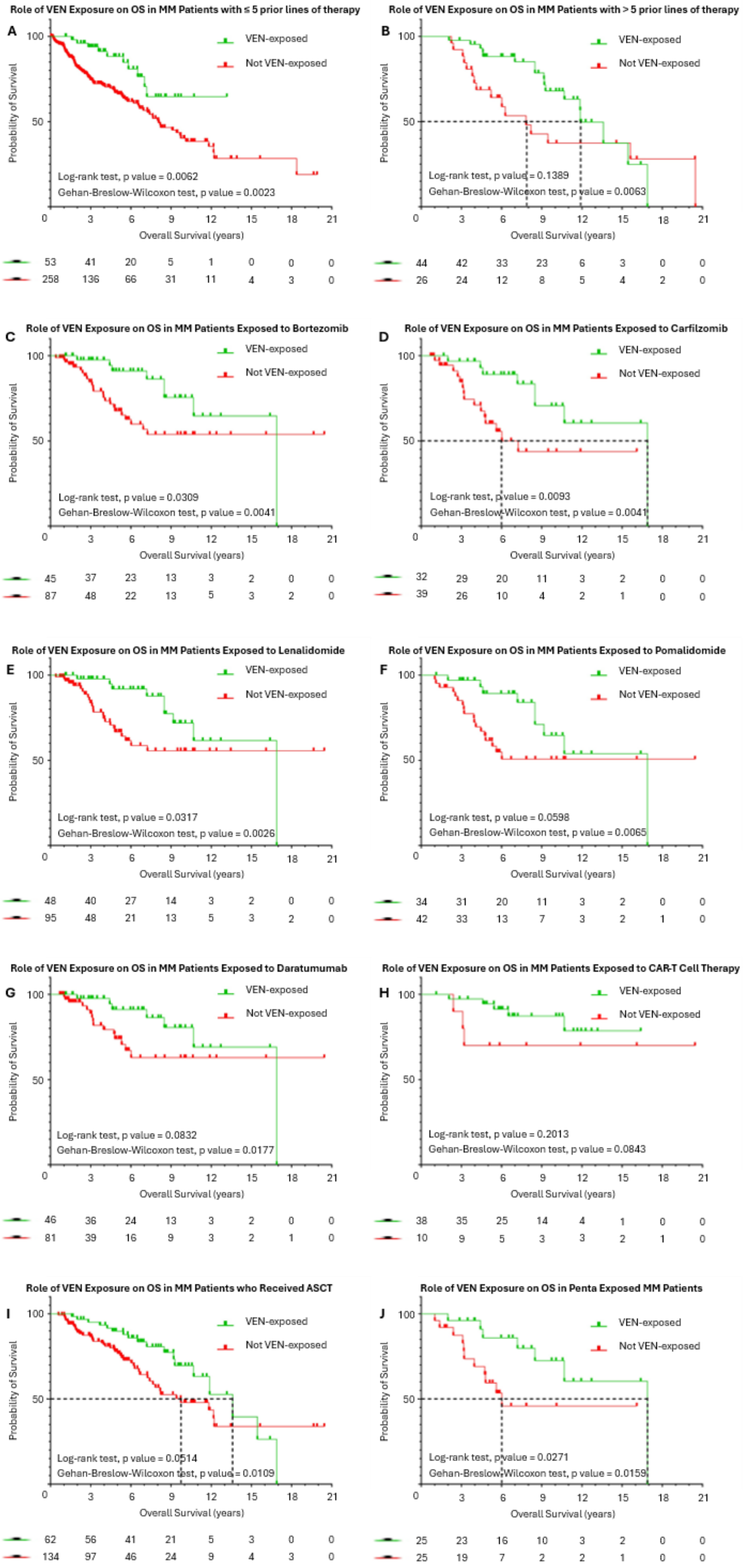

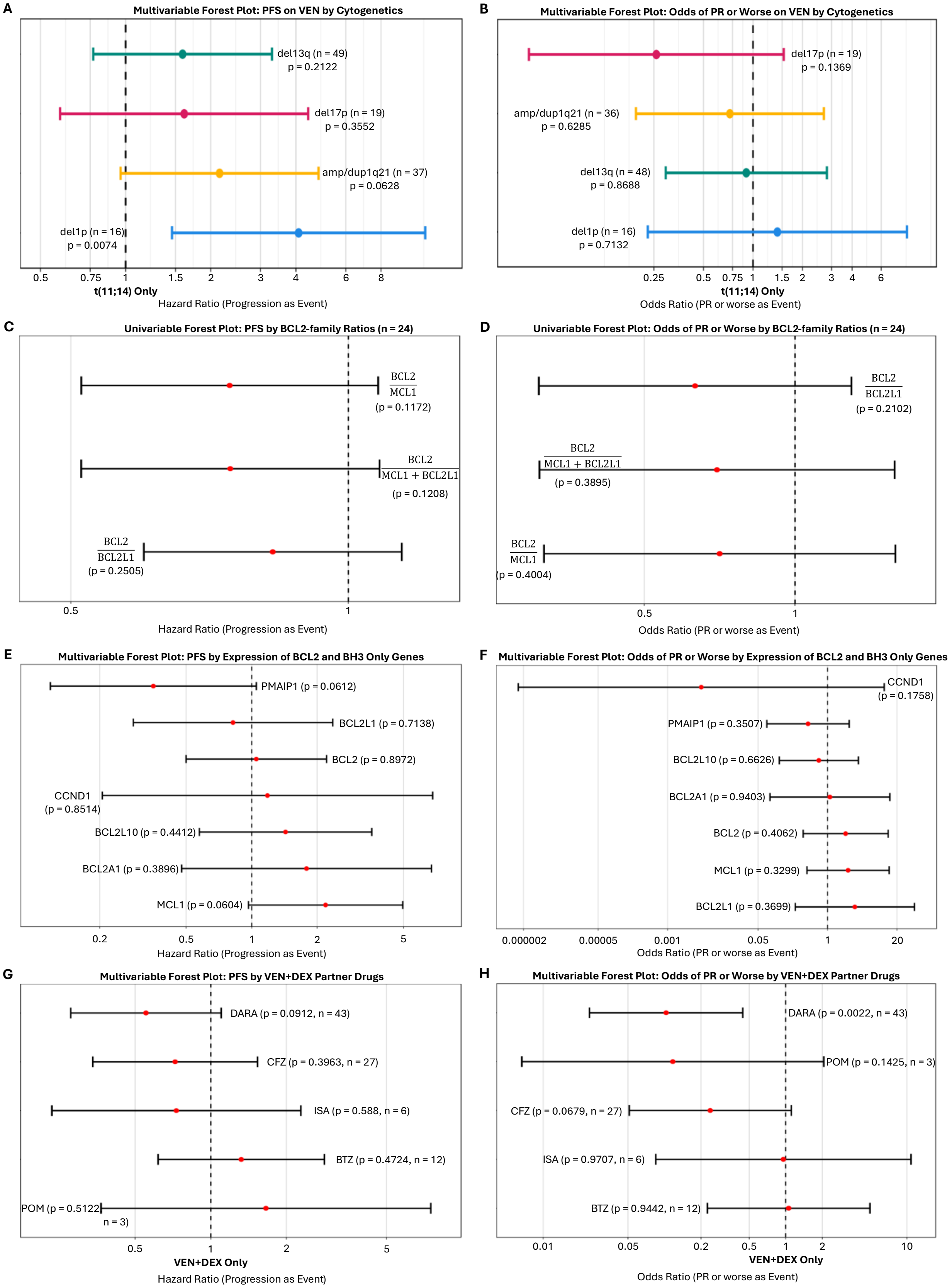


**Supplementary Fig. 2** Forest plots featuring Cox proportional hazards for progression as the event and logistic regression analyses for depth of response, PR or worse, as the event across cytogenetics using multivariable analyses **(A)** & **(B)**, BCL2-family ratios using univariable analyses **(C)** & **(D)**, BCL2-family and BH3-only gene expression using multivariable analyses **(E)** & **(F)**, and VEN partner drugs using multivariable analyses **(G)** & **(H)**.

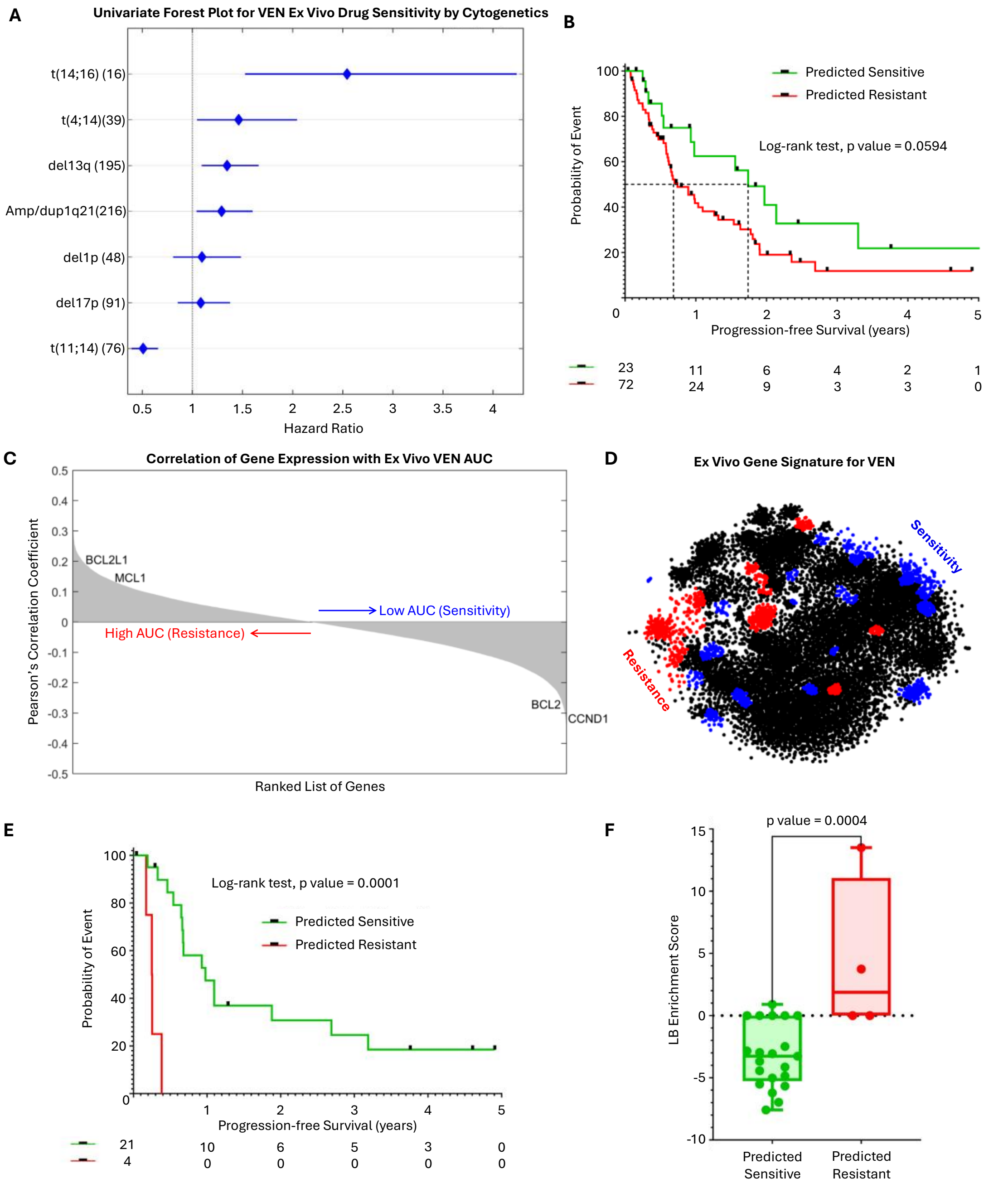


**Supplementary Fig. 3** Ex vivo analysis of primary MM samples identifies transcriptomic signatures of single agent VEN sensitivity in t(11;14) MM. **(A)** A univariable forest plot for VEN ex vivo sensitivity (shown as a hazard ratio estimated from percent cells survived) for each cytogenetic abnormality tested. **(B)** A Kaplan-Meier comparison of PFS on VEN between MM patients predicted to be sensitive to VEN using cytogenetics as input to a regression tree model trained on paired cytogenetics and ex vivo AUC. **(C)** A waterfall plot of ranked genes from positively correlated to negatively correlated expression with ex vivo response. **(D)** An MM transcriptomic map showing significantly enriched gene clusters for ex vivo repose to VEN. **(E)** A Kaplan-Meier survival comparison of PFS between MM patients treated with VEN predicted to be sensitive vs resistant using the regression tree model. **(F)** A boxplot comparing the ssGSEA enrichment scores for LB using an unpaired t-test between predicted sensitive and resistant patients by the regression tree model.

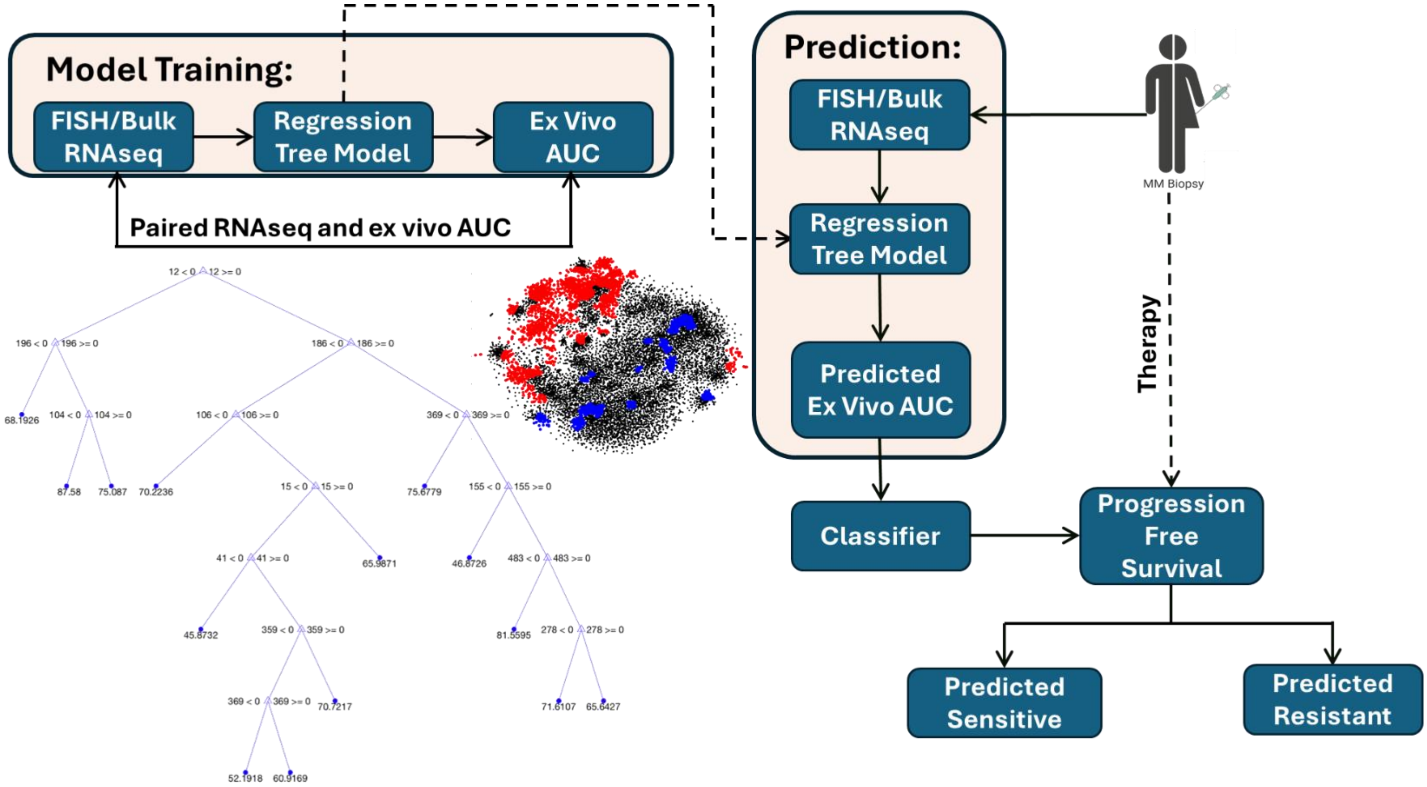


**Supplementary Fig. 4** A graphical representation of the regression tree model training and implementation to identify sensitive and resistant patients to a given drug.

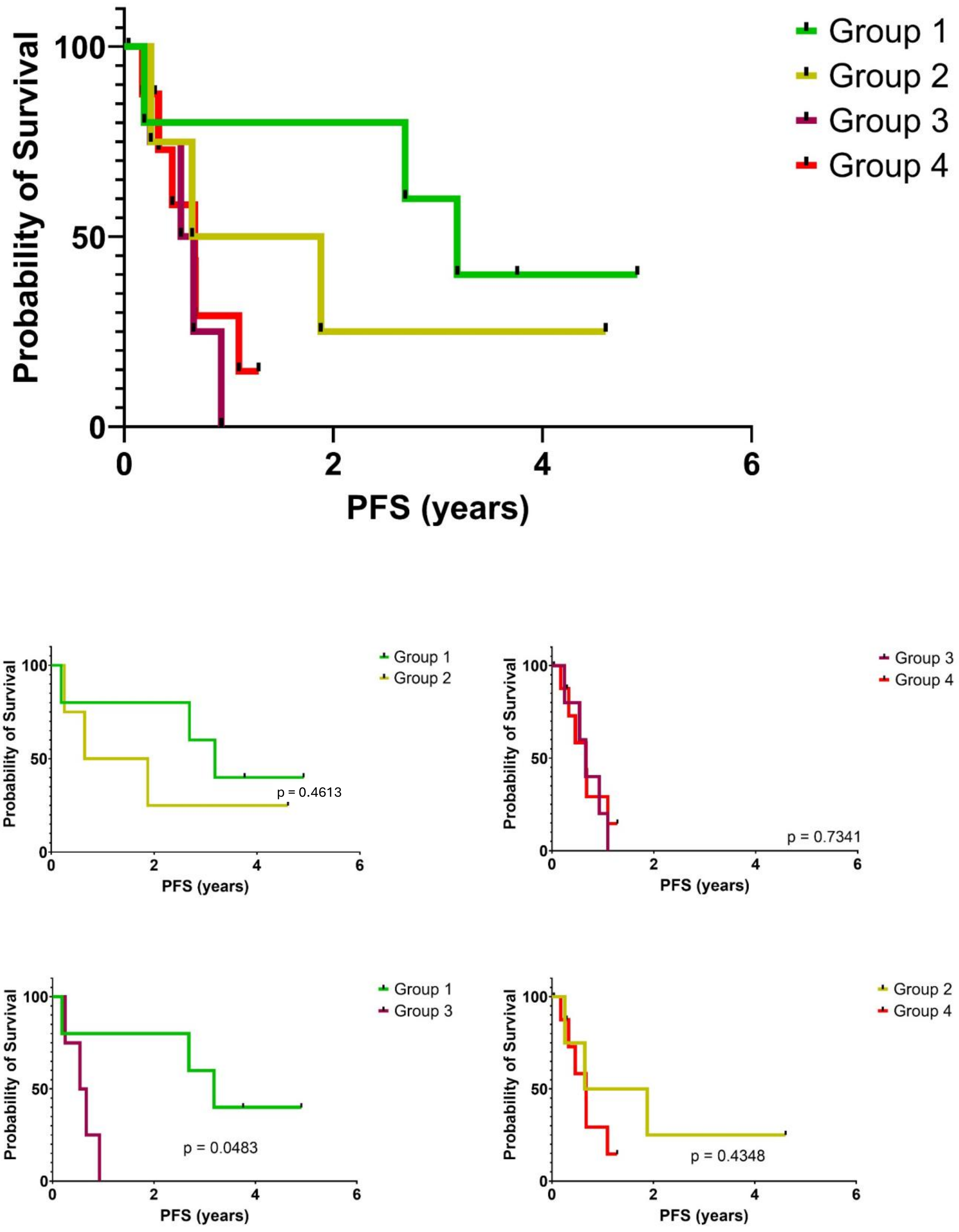


**Supplementary Fig. 5** Kaplan-Meier Survival Plots comparing the VEN PFS of patients between Groups 1 to 4.

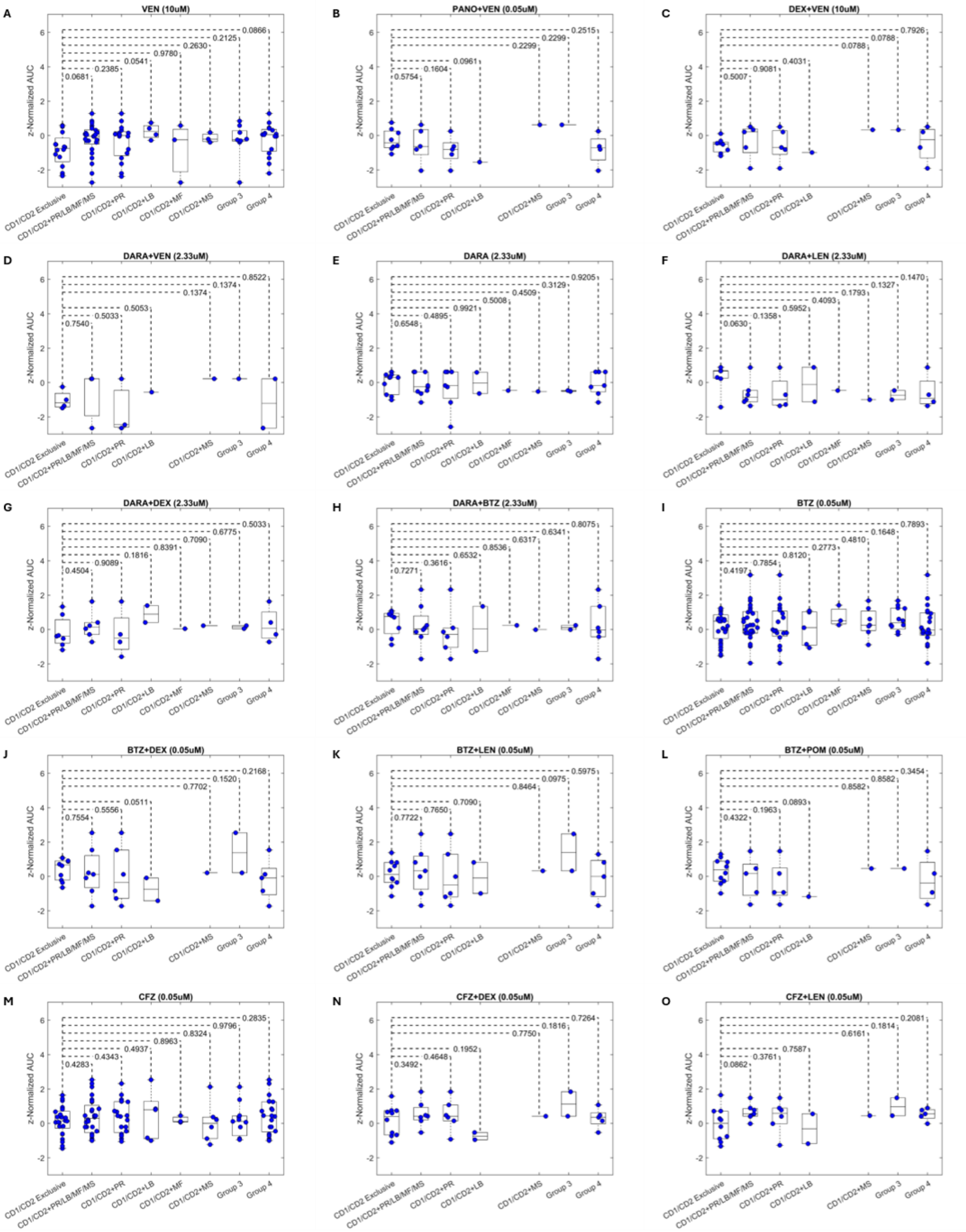


**Supplementary Fig. 6** Box plots comparing CD1/CD2 exclusive group with other functionally complex groups using unpaired t-tests for SOC single agents/combinations, and experimental agents.

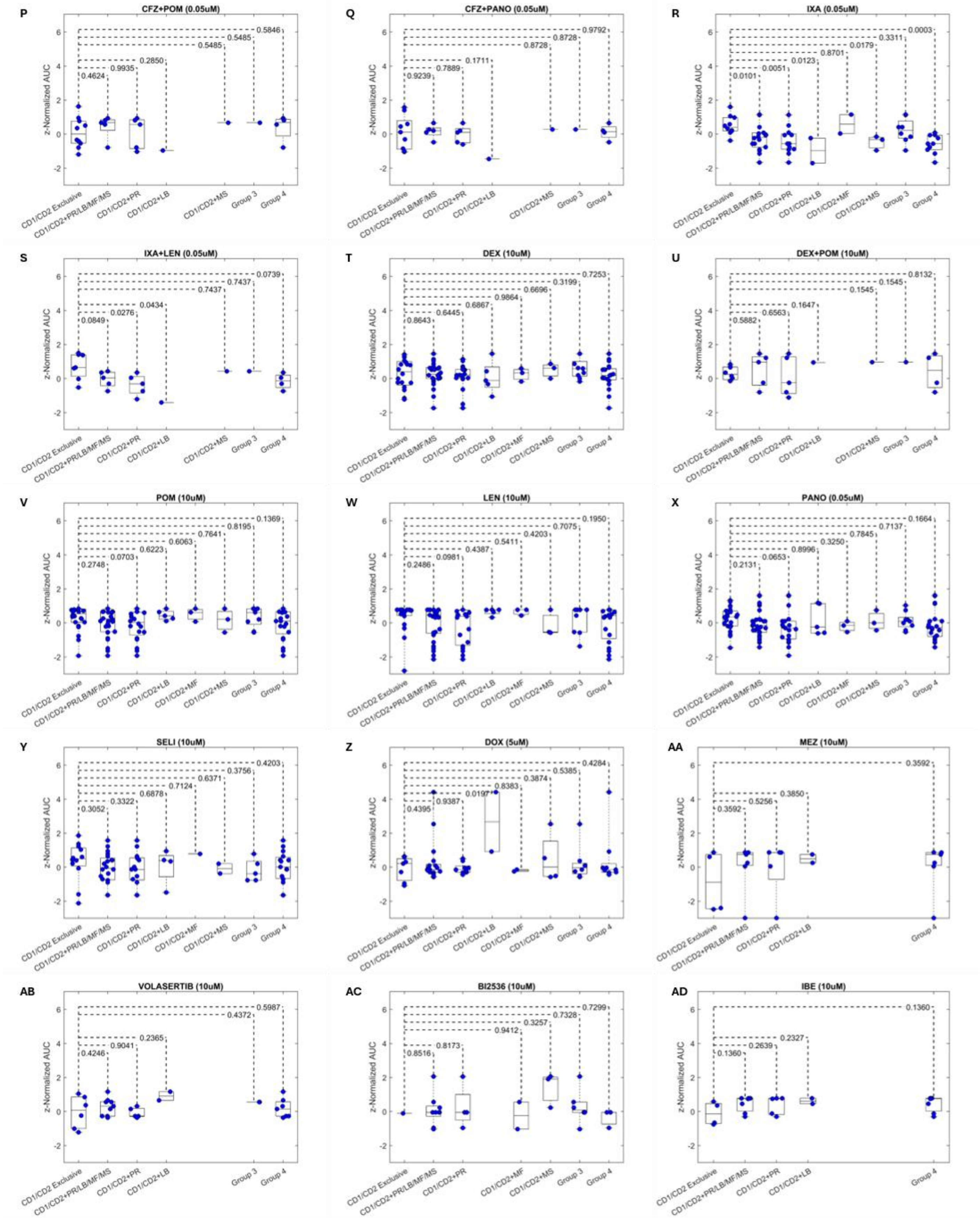


**Supplementary Fig. 6 (continued)** Box plots comparing CD1/CD2 exclusive group with other functionally complex groups using unpaired t-tests for SOC single agents/combinations, and experimental agents.

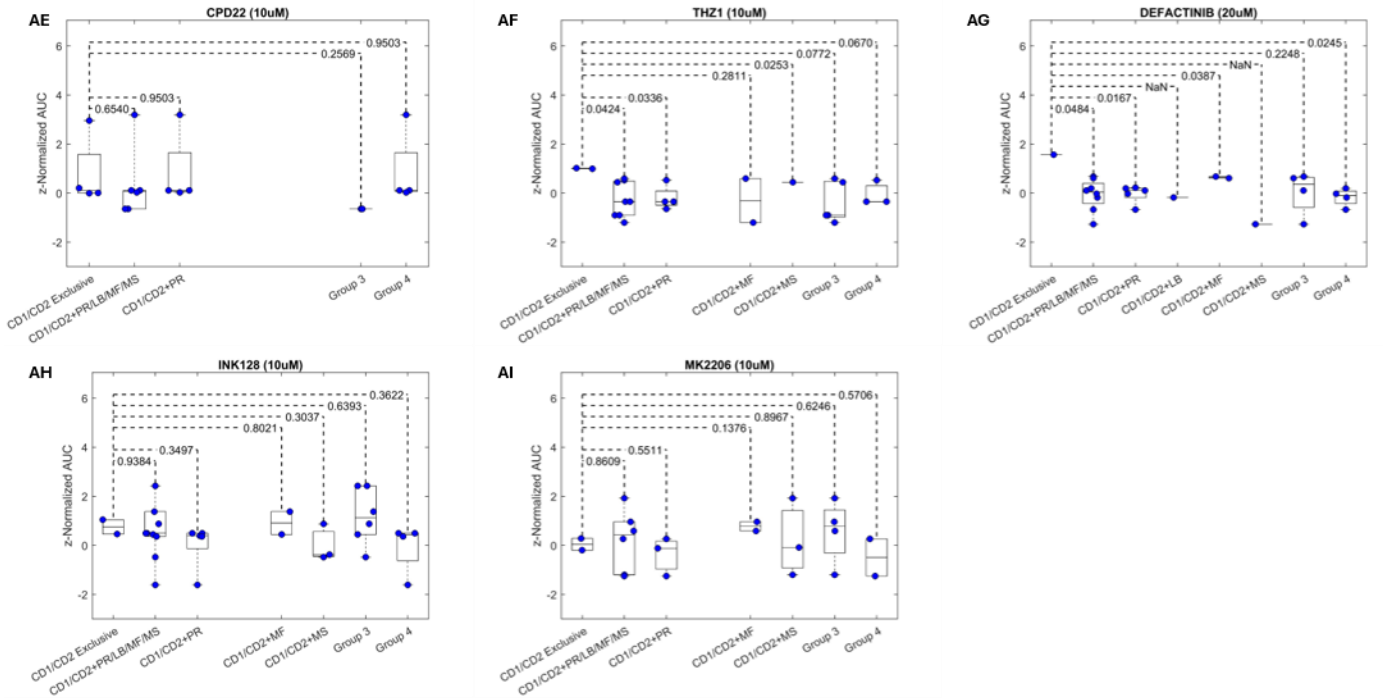


**Supplementary Fig. 6 (continued)** Box plots comparing CD1/CD2 exclusive group with other functionally complex groups using unpaired t-tests for SOC single agents/combinations, and experimental agents.

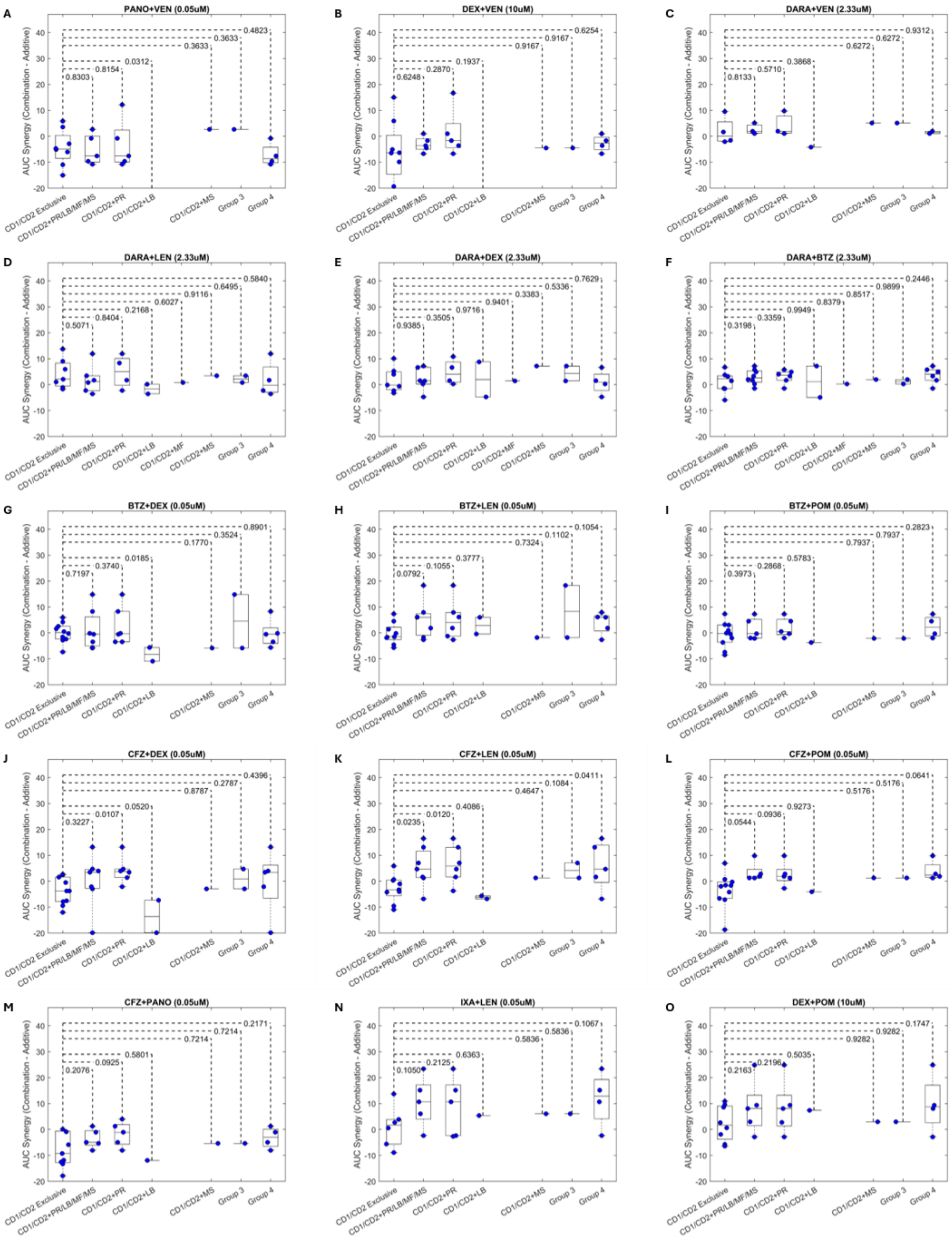


**Supplementary Fig. 7** Box plots comparing CD1/CD2 exclusive group with other functionally complex groups using unpaired t-tests for each two-drug combination by ex vivo synergy.
